# The Interplay of Perceived Mental Health and Suicidal Ideation among Aboriginal Peoples in Canada: A Secondary Analysis of Aboriginal Peoples Survey

**DOI:** 10.1101/2024.09.28.24314555

**Authors:** Olasehinde Ben Adebayo

**Affiliations:** College of Health Professions, Bellarmine University, Louisville, KY, USA

**Keywords:** Suicide Ideation, Mental Health, Aboriginal Peoples, Socioeconomic Status

## Abstract

The disproportionately high suicide rates among Aboriginal populations in Canada, linked to historical trauma and systemic inequities, necessitate targeted mental health interventions. This study investigated the relationship between perceived mental health and suicidal ideation among the Aboriginal population in Canada, First Nations, Inuit, and Métis, using data from the 2012 Aboriginal Peoples Survey (APS) (n=4,725). The analysis employed chi-square tests and bivariate logistic regression to examine the association between self-perceived mental health, suicidal ideation, and socio-demographic factors. Results revealed significant associations between suicidal ideation, age group (p<.001), and sex (p<.001), with younger individuals (18-24 years) and females reporting a higher prevalence of suicidal thoughts. Also, bivariate logistic regression showed that being between the ages of 25 and 34 was a strong predictor of suicidal thoughts (OR = 1.954, p =.000), while being female (OR =.648, p =.000) and having a higher socioeconomic status (OR =.678, p =.000) were weaker predictors. Although Aboriginal identity was not significantly associated with suicidal ideation, self-perceived mental health was. These results made it clear that focused and targeted mental health interventions and culturally appropriate suicide prevention strategies are needed right away in Aboriginal communities in Canada. These strategies need to meet the needs of vulnerable groups, like younger people and women.

## Introduction

Between 2011 and 2016 in Canada, suicide rates were noticeably higher among Aboriginal populations in comparison to non-Aboriginal populations (Kumar & Tjepkema, 2019). The disproportionately high rates of suicide among the Canadian Aboriginal population, comprising the First Nations, Inuit, and Métis populations, compared to the non-Aboriginal population necessitate a deeper understanding of the underlying factors contributing to this mental health crisis. Existing research has revealed a significant burden of mental health problems and suicides among Aboriginal Canadians (Kral, 2016; Nelson & Wilson, 2017) with First Nations and Inuit youth experiencing particularly alarming suicide rates (Nelson & Wilson, 2017; Orkin et al., 2013). These disparities are often attributed to a complex interplay of socio-economic, cultural, and historical factors, which include poverty, discrimination, and the enduring effects of colonisation and residential school experiences (Kral, 2016; Toombs et al., 2023; Wilk et al., 2017). The economic disadvantages faced by many Indigenous communities, coupled with historical trauma and systemic racism, can significantly impact mental health status and increase the likelihood of suicidal ideation (Kral, 2016; Pandeya et al., 2021; Smye et al., 2023), making it a crucial area for investigation.

This study examines the relationship between perceived mental health and suicidal ideation within these Aboriginal populations through a secondary data analysis. This study aims to determine the extent to which perceived poor mental health is associated with suicidal ideation among First Nations, Inuit, and Métis populations in Canada. By addressing the research question, “To what extent does perceived poor mental health correlate with suicidal thinking among First Nations, Inuit, and Métis populations in Canada?”, this study seeks to identify a critical intervention point for targeted mental health programmes and suicide prevention efforts. This study, by elucidating the relationship between perceived mental health and suicidal thoughts within these communities, can inform the development of culturally sensitive mental health interventions and suicide prevention strategies tailored to the specific needs of Aboriginal peoples in Canada.

## Methodology

### Participants

The secondary data for this research was derived from the 2012 Aboriginal Peoples Survey (APS), a comprehensive survey conducted by Statistics Canada (Government of Canada, 2023). The Aboriginal Peoples are defined as the First Nations, Inuit, and Métis people. The Aboriginal people surveyed are 18 years of age and older. The sample consisted of 55.2% females and 44.8% males. 39.3%) of the participants identified as First Nations, 36.1% as Métis and 24.6% as Inuit (Government of Canada, 2023). The largest age group in the study was young adults aged 18 to 24 years (31.7%) (Government of Canada, 2023)

### Procedure

This study used secondary data analysis methods to analyze existing APS data. The secondary data in APS is cost-effective, timesaving, and contains a wider range of information. The APS data was selected to ensure reliable and valid responses. The survey included standardised questions across various domains, including health, education, employment, and cultural participation. The following variables of interest were extracted from the APS dataset for this research: perceived mental health, suicidal thoughts, and socioeconomic status (SES).

The analysis was conducted using data obtained from Statistics Canada’s official repository at https://www150.statcan.gc.ca/n1/tbl/csv/41100011-eng.zip (Government of Canada, 2023). Extensive data cleaning was performed using R-Studio 4.1.2 due to the initial disarray of the dataset. Subsequently, data analysis was carried out using SPSS version 22 (Appendix A). Descriptive statistics were used to examine perceived mental health, suicidal thoughts, and SES across different Aboriginal identity groups. To investigate the association between sociodemographic variables and suicidal ideation and self-perceived mental health among Aboriginal people in Canada. A chi-square cross-tabulation analysis was performed to identify potential relationships between categorical variables due to its suitability and ability to identify relationships between categorical variables (Momeni et al., 2017). The researcher used the chi-square to test or identify the relationship between the categorical variables (suicide ideation and perceived mental health) and the socio-demographic variables (Aboriginal identity, sex, and age group).

Furthermore, bivariate logistic regression analysis was employed to examine the association between perceived mental health and suicidal ideation among Aboriginal people in Canada. This method was selected as logistic regression is well-suited for modelling binary outcomes (Starbuck, 2023). The presence or absence of suicidal ideation is a dichotomous variable. The researcher calculated and utilized odds ratios (OR) to understand the strength and direction of the relationship between the outcomes. I conducted all statistical analyses using SPSS software (Appendix A) to ensure robust and reliable results.

### Data Preparation

The dataset underwent filtering, recoding, and transformation processes to maintain data integrity and relevance to the study’s focus on Aboriginal identity, socioeconomic status (SES), and mental health outcomes, particularly suicidal thoughts. These steps were taken to convert the raw data into a suitable format for statistical analysis. The data required extensive cleaning due to its messy state. The dataset was filtered to include only relevant categories for Aboriginal identity (First Nations, Métis, and Inuit). Irrelevant categories were removed, such as adult age cohorts not pertinent to the study’s socioeconomic and mental health indicators, specifically those below 18 years of age. The “Both sexes” category was also eliminated, focusing on males and females separately.

A crucial step in the data-cleaning process involved creating a variable indicating suicidal ideation. This was achieved by recording responses from the self-perceived mental health and suicidal thoughts Likert measures. The researcher coded 1 (yes) for participants who selected responses indicating suicidal ideation, and 0 (no) for others. This resulted in a dichotomous variable named “Suicidal ideation,” which served as the dependent variable in subsequent analyses. Lastly, missing observations were removed to ensure data reliability for analysis.

## Measures

### Perceived Mental Health

The overall mental health of individuals was assessed using a self-report question in the APS. Responses ranged from “excellent” to “poor,” providing a subjective measure of perceived mental well-being. For analysis, perceived mental health was dichotomized into “good” (combining excellent/very good/good) and “poor” (combining fair/poor).

### Suicidal Thoughts

Suicidal thoughts were assessed by inquiring whether respondents had any thoughts of suicide within the past year. This dichotomous variable (yes/no) represents the presence or absence of suicidal thoughts and served as the outcome variable in the logistic regression analysis.

### Socioeconomic Status (SES)

Socioeconomic status was determined based on reported income and education levels in the APS dataset. The education level was determined based on the highest grade completed. The analysis included SES as a covariate to control for its potential confounding effect on the relationship between perceived mental health and suicidal thoughts.

### Sociodemographic Characteristics

Age, gender, and Aboriginal identity were included as socio-demographic variables in the survey. Age groups were consolidated to include those 18 years and older to assess changes in mental health attitudes across different life stages. Both sexes were included to identify potential gender-specific differences in mental health outcomes. The Aboriginal identity variable comprised three categories: First Nations, Métis, and Inuit

## Results

The initial analysis of the dataset presented in Table 1 shows the frequency distribution of the variables used in the study. The results indicate that 39.3% of the respondents identified as First Nation (n = 1858), 24.6% as Inuit (n = 1160), and 36.1% as Métis (n = 1707). Similarly, the result shows that among the age groups, 31.7% of the respondents (n = 1496) were aged 18 to 24 years, followed by individuals aged between 45 and 54 years, constituting 23.0% of the sample (n = 1089), individuals aged 35 to 44 years constituted 22.7% (n = 1072), and 22.6% (n = 1068) were those aged 25 to 34 years. The result further shows the sample was slightly skewed towards females (55.2%) while 44.8% (n = 2117) were males. The age group with the highest representation was 18-24 years (31.7%), indicating a focus on younger individuals. A significant portion of the sample (39.2%) reported having experienced suicidal thoughts.

**Table 1:**
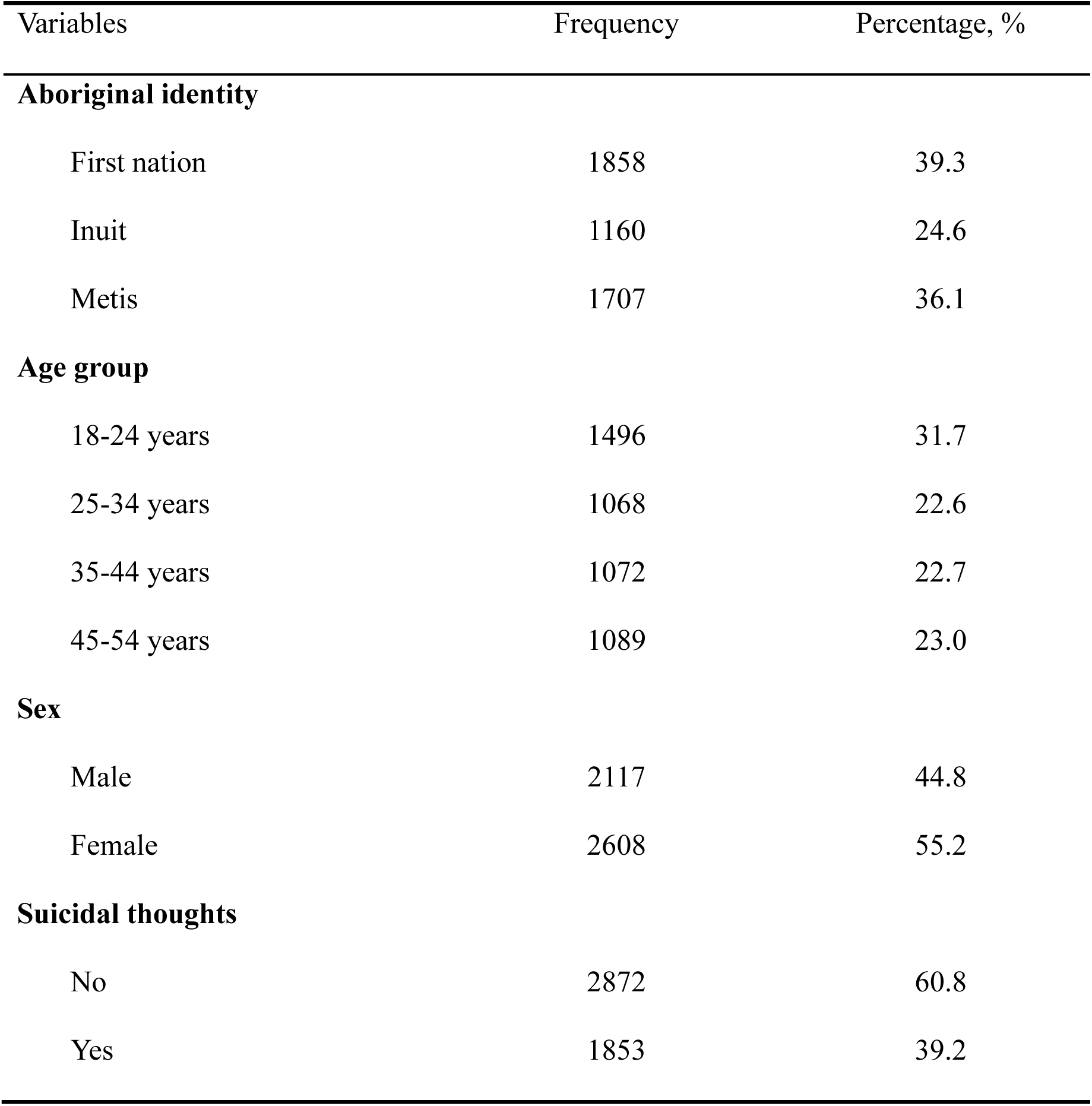
Frequency distribution of the variables used in the study.

| Variables | Frequency | Percentage, % |
| --- | --- | --- |
| **Aboriginal identity** |  |  |
| First nation | 1858 | 39.3 |
| Inuit | 1160 | 24.6 |
| Metis | 1707 | 36.1 |
| **Age group** |  |  |
| 18-24 years | 1496 | 31.7 |
| 25-34 years | 1068 | 22.6 |
| 35-44 years | 1072 | 22.7 |
| 45-54 years | 1089 | 23.0 |
| **Sex** |  |  |
| Male | 2117 | 44.8 |
| Female | 2608 | 55.2 |
| **Suicidal thoughts** |  |  |
| No | 2872 | 60.8 |
| Yes | 1853 | 39.2 |

### Chi-Square Test of Association (Suicide Ideation)

Table 2 presents the cross-tabulation results of socio-demographic variables associated with suicide ideation among Aboriginal people in Canada. Of the respondents, 39.7% of the first nation individuals indicated they have experienced suicide ideation in the past 12 months, 40.1% of the Inuit individuals have experienced suicide thoughts, 38.1% of the Metis individuals have experienced suicide thoughts in the past 12 months, and 60.3%, 59.9% and 61.9% respectively, indicated they did not. There was no significant association between suicide ideation and aboriginal identity (p = .507). Individuals aged between 18 and 24 years had a higher prevalence of suicide ideation (50.3%) compared to other age groups. The result shows that there is a statistically significant association between suicide ideation and age group (p = .000). Females had a higher prevalence of suicide ideation than males, at 43.5% versus 34.0%. Among males, 66.0% reported no suicide ideation, while among females, 56.5% reported no suicide ideation. The result also shows a significant association between gender and suicide ideation (p = .000).

**Table 2:** Cross-tabulations of socio-demographic variables with suicide ideation in past years among Aboriginal people in Canada.

|  | Suicide ideation, % |  | p-value |
| --- | --- | --- | --- |
|  | No | Yes |  |
| **Aboriginal identity** |  |  | **0.507** |
| First nation | 60.3 | 39.7 |  |
| Inuit | 59.9 | 40.1 |  |
| Metis | 61.9 | 38.1 |  |
| **Age group** |  |  | **0.000** |
| 18-24 years | 49.7 | 50.3 |  |
| 25-34 years | 65.3 | 34.7 |  |
| 35-44 years | 66.6 | 33.4 |  |
| 45-54 years | 65.9 | 34.1 |  |
| **Sex** |  |  | **0.000** |
| Male | 66.0 | 34.0 |  |
| Female | 56.5 | 43.5 |  |

$p < .001$

All the socio-demographic characteristics variables were statistically associated with self-perceived mental health (p < .001). Among the age groups, the results were highly significant (p < .001), with younger age groups (18-24 years) showing higher percentages of serious suicidal thoughts in the past 12 months compared to older age groups. This suggests that younger individuals experience significant changes such as puberty, life expectations, and social pressure, which can contribute to stress and anxiety Backes et al., 2024)

Furthermore, there was a significant association between sex, self-perceived mental health, and suicidal thoughts (p < .001). A higher percentage of males reported excellent or very good self-perceived mental health (48.7%), while females reported higher percentages of fair or poor self-perceived mental health and suicidal thoughts (65.4%). The researcher found a significant association between Aboriginal identity and self-perceived mental health (p =.032). A significant association was found between age group and self-perceived mental health (p < .001). Younger individuals (18-24 years) reported higher rates of poor mental health and suicidal thoughts. A significant association was found between sex and self-perceived mental health (p < .001). Females reported higher rates of fair or poor mental health and suicidal thoughts compared to males.

### Bivariate Logistic Regression

The result of the bivariate logistic regression model examining the relationship between perceived mental health and suicidal thoughts is presented in Table 4. The result shows that individuals with poor perceived mental health have a high likelihood of contemplating suicidal thoughts, although this was not statistically significant (OR = 1.019, p = 0.248). This suggests that the likelihood of contemplating suicidal thoughts was not meaningfully different across varying levels of perceived mental health.

**Table 3:**
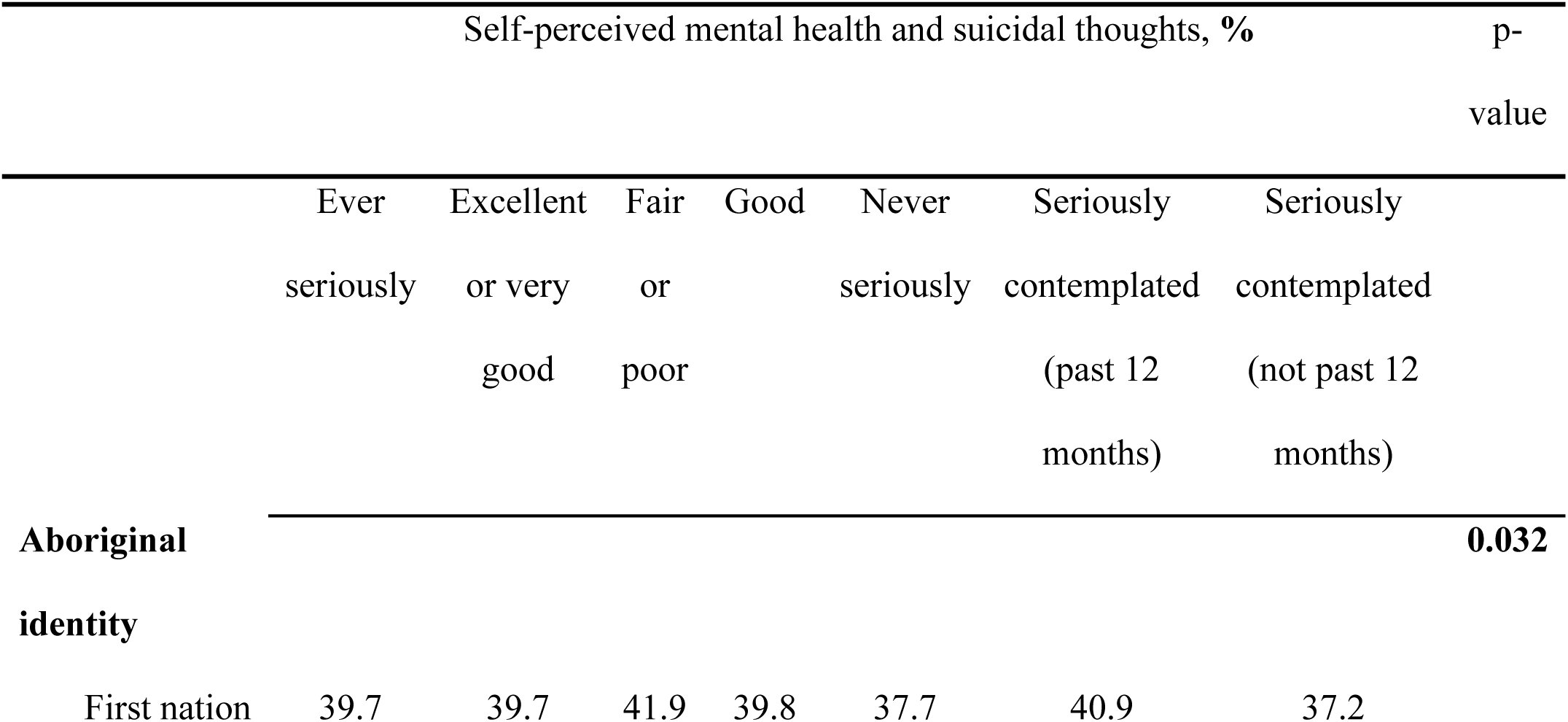
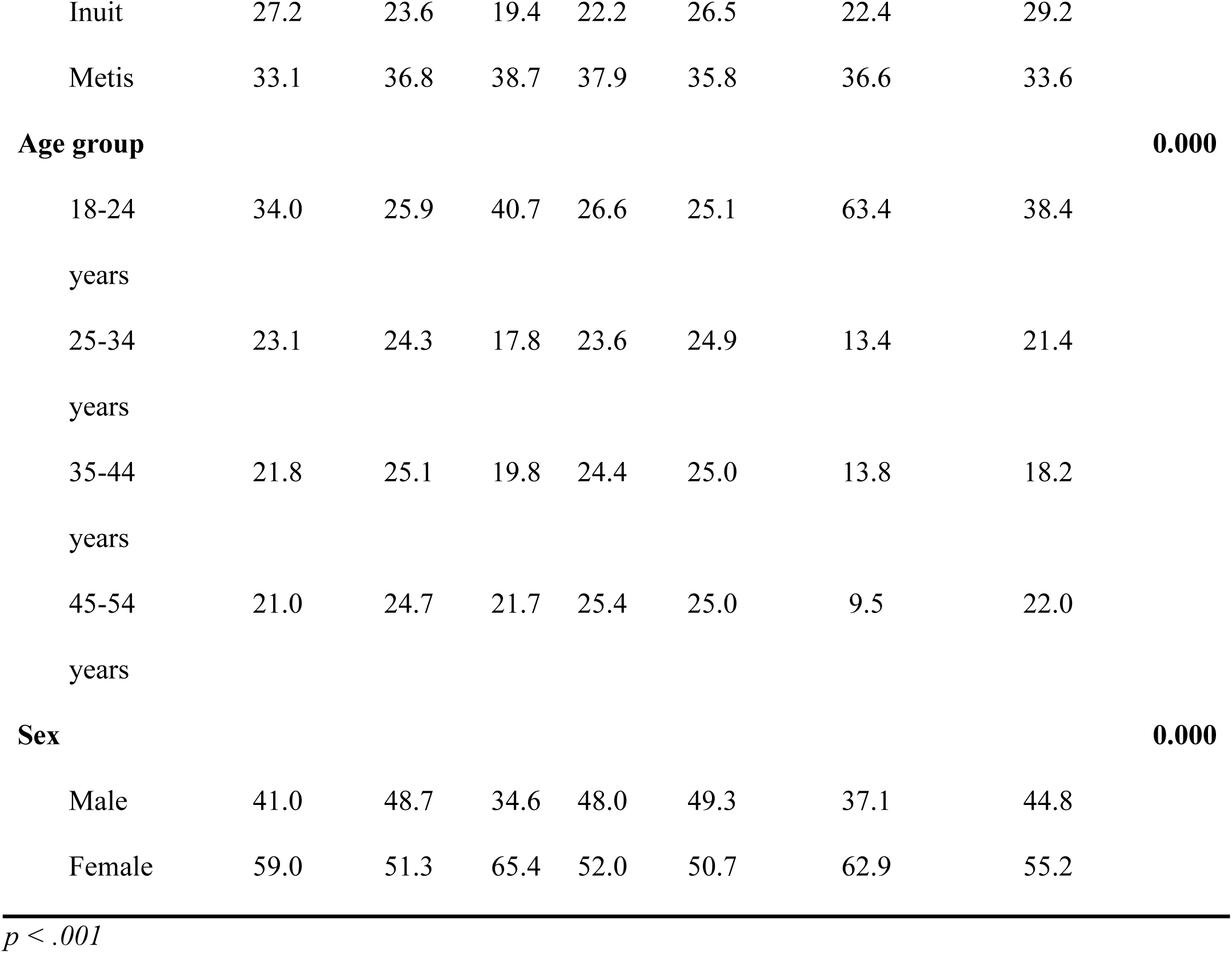
Cross Tabulations of Socio-demographic variables with perceived mental health in past years among Aboriginal people in Canada (self-perceived mental health)

**Table 4:** Bivariate Logistic Regression model: A two-variable logistic regression model was used to look at the link between how mentally healthy someone thinks they are and suicidal thoughts while considering sociodemographic factors.

| Variables included | B | Wald | p-value | Odds ratio<br>(OR) | 95% CI for OR |  |
| --- | --- | --- | --- | --- | --- | --- |
|  |  |  |  |  | Lower | Upper |
| **Constant** | -.609 | 36.629 | .000 | .544 |  |  |
| **Perceived mental health**<br>**and suicidal** | .019 | 1.337 | .248 | 1.019 | .987 | 1.053 |
| **Aboriginal identity** |  |  |  |  |  |  |
| <sup>1</sup>First nation |  | .938 | .626 |  |  |  |
| Inuit | .067 | .895 | .344 | 1.069 | .931 | 1.227 |
| Metis | .020 | .064 | .800 | 1.020 | .872 | 1.194 |
| **Age group** |  |  |  |  |  |  |
| 18-24 years |  | 109.713 | .000 |  |  |  |
| 25-34 years | .670 | 64.896 | .000 | 1.954 | 1.660 | 2.300 |
| 35-44 years | .032 | .121 | .728 | 1.032 | .863 | 1.235 |
| 45-54 years | -.043 | .215 | .643 | .958 | .800 | 1.147 |
| **Sex** |  |  |  |  |  |  |
| Male |  |  |  |  |  |  |
| Female | -.434 | 49.454 | .000 | .648 | .574 | .731 |
| **Socioeconomic status** | -.389 | 29.894 | .000 | .678 | .589 | .779 |

$R^2 = 0.058$ (Nagelkerke), $0.043$ (Cox & Snell). Model $\chi^2_8 = 65.353$ , $p = 0.000$ .
Reference categories : Aboriginal identity – First Nation ;
Age group – 18-24 years;
Sex: Male

Regarding Aboriginal identity, the odds ratios for Inuit (OR = 1.069, p = 0.344) and Métis (OR = 1.020, p = 0.800) were not statistically significant, meaning that Inuit and Métis individuals were not more likely to experience suicidal thoughts compared to First Nations individuals. The result suggests that the odds of experiencing suicidal thoughts were essentially the same across these Aboriginal identities.

Among the age groups, individuals between the ages of 25 and 34 (OR = 1.954, p = 0.00) are more likely to experience suicidal thoughts than others. This implies that an individual between the ages of 25 and 34 is associated with a 1.954 increase in the odds of being involved in suicidal ideations. Furthermore, the result shows that females have a lower odd of experiencing suicidal thoughts than males (OR = 0.648, p = 0.000). Lastly, socioeconomic status also showed a significant negative association with suicidal thoughts (OR = 0.678, p = 0.000), suggesting that higher socioeconomic status (reported income and education levels) is associated with lower odds of experiencing suicidal thoughts. The model showed a net improvement in classification, with the percentage of correct classifications increasing from the null model to the fitted model. This indicates that the inclusion of perceived mental health, Aboriginal identity, age group, sex, and socioeconomic status as predictors improved the model’s ability to correctly classify individuals with and without suicidal thoughts

The model fit statistics show that the model is a moderately good fit for the data (Nagelkerke’s R² = 0.058 and Cox & Snell’s R² = 0.043). The model chi-square test is statistically significant (χ² = 65.353, p < 0.001), indicating that the model significantly predicts the outcome variable.

## 4. Discussion

Findings from the study showed that there was no significant association between suicide ideation and Aboriginal identity. Furthermore, there was a statistically significant association between suicide ideation and age group, as well as a significant association between gender and suicide ideation. This aligns with the findings of Zhang et al., who reported a 60% increase in female suicide rates over male suicide rates in their study on gender differences in suicidal ideation and health risk behaviour (Zhang et al., 2019). However, this contradicts the findings of Lemstra et al. (2013), who found no significant influence of age or gender on the likelihood of suicide ideation. Their study revealed that suicidal ideation disproportionately affected girls, with 27.8% reporting such thoughts among youths in Canada, compared to 15.2% for boys, a finding that aligns with my findings.

The data suggests that age and sex are significant factors influencing suicide ideation and self-perceived mental health among Aboriginal people in Canada. Younger individuals and females appear to be particularly vulnerable groups. While Aboriginal identity was not significantly associated with suicide ideation in Table 2, it was significantly associated with self-perceived mental health in Table 3, suggesting a more nuanced relationship. The findings further showed that individuals with poor perceived mental health have a high likelihood of contemplating suicidal thoughts, although this was not statistically significant. This agrees with the findings of Akram et al. (2020) which analysed the link between suicidal ideation and psychiatric symptoms like depression, mania, psychosis, and stress in UK university students. The result concluded that suicidal ideation was significantly associated with symptoms of depression, mania, psychosis, and stress (Akram et al., 2020).

Among Aboriginal identities, being an Inuit individual is associated with an increase in the odds of experiencing suicidal thoughts compared to non-Inuit individuals. Similarly, there are higher odds of Metis individuals experiencing suicidal thoughts than non-Metis individuals. The lack of a significant association between perceived mental health and suicidal thoughts indicates that interventions should not solely focus on improving perceived mental health but should also address broader sociodemographic factors such as occupational status, sexual orientation, educational level, immigration status, and family structure. This study has several limitations. The use of secondary data analysis restricts the analysis to the variables available within the APS, potentially omitting other important factors Finally, stigma, fear of judgment, or memory errors may lead to underreporting or overreporting, compromising the accuracy of self-reported mental health data.

## Ethical Considerations

### Ethical Statement

The submitted manuscript did not involve human or animal research. This study utilised secondary data. No ethical approval was sought or required for this study. It is a research review submitted in Partial fulfilment of the requirement for PhD Health Professions Education, 2024SU Statistics in Health Professions Education II (HLTH-862-ON)/Summer 2024 at Bellarmine University Kentucky, United States of America.

## Conclusions

This analysis reveals the complex interplay of perceived mental health, suicidal ideation, and socio-demographic factors among Aboriginal peoples in Canada. The findings from the secondary data used accentuate the heightened susceptibility of younger individuals, particularly those aged 25-34 years old and females, to suicidal thoughts. This emphasises the need for targeted mental health interventions and culturally sensitive suicide prevention strategies for the population. While Aboriginal identity alone may not directly predict suicidal ideation, its significant association with self-perceived mental health points to a subtle interplay that warrants further investigation.

The research utilised secondary data, meaning the researcher did not participate in the data collection process. This lack of control raises concerns regarding potential inconsistencies, errors, or biases within the data, which may affect the quality, reliability, and validity of the findings. Additionally, the reliance on self-reported data highlights the need for future research to employ prospective designs that incorporate a wider array of variables, including cultural determinants and historical trauma. This approach would facilitate a more comprehensive understanding of the multifaceted factors influencing mental well-being and suicidal ideation among Aboriginal communities in Canada. To improve understanding, future investigations should prioritize prospective research designs to elucidate causal relationships between the variables examined herein and those identified for future inquiry. Additionally, exploring the impact of cultural determinants, such as perceived stress, religious affiliation, and historical trauma, on mental well-being and suicidal ideation within these populations is warranted. By addressing these knowledge gaps, researchers and policymakers can collaboratively develop more effective, culturally sensitive interventions to alleviate the mental health crisis and reduce suicide rates within these populations.

## Data Availability

All data produced are available online at https://www150.statcan.gc.ca/n1/tbl/csv/41100011-eng.zip

## APPENDIX A

SPSS Output File

CROSSTABS

/TABLES=Self_perceived_mental_health_and_suicidal_thoughts BY Aboriginal_identity

Age_group Sex

/FORMAT=AVALUE TABLES

/STATISTICS=CHISQ CORR

/CELLS=COUNT ROW COLUMN

/COUNT ROUND CELL.

**Crosstabs**

**Self_perceived_mental_health_and_suicidal_thoughts * Aboriginal_identity**

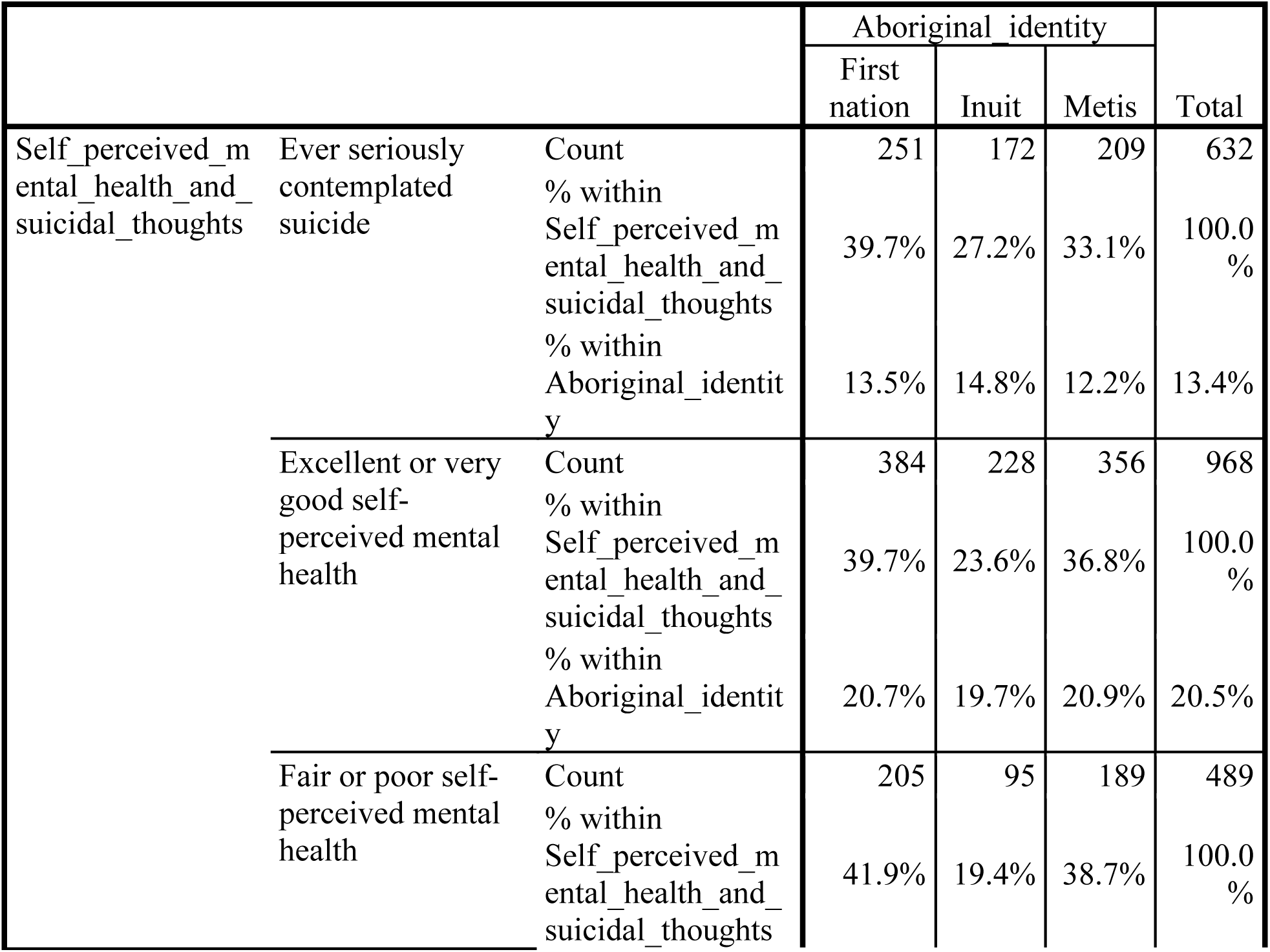
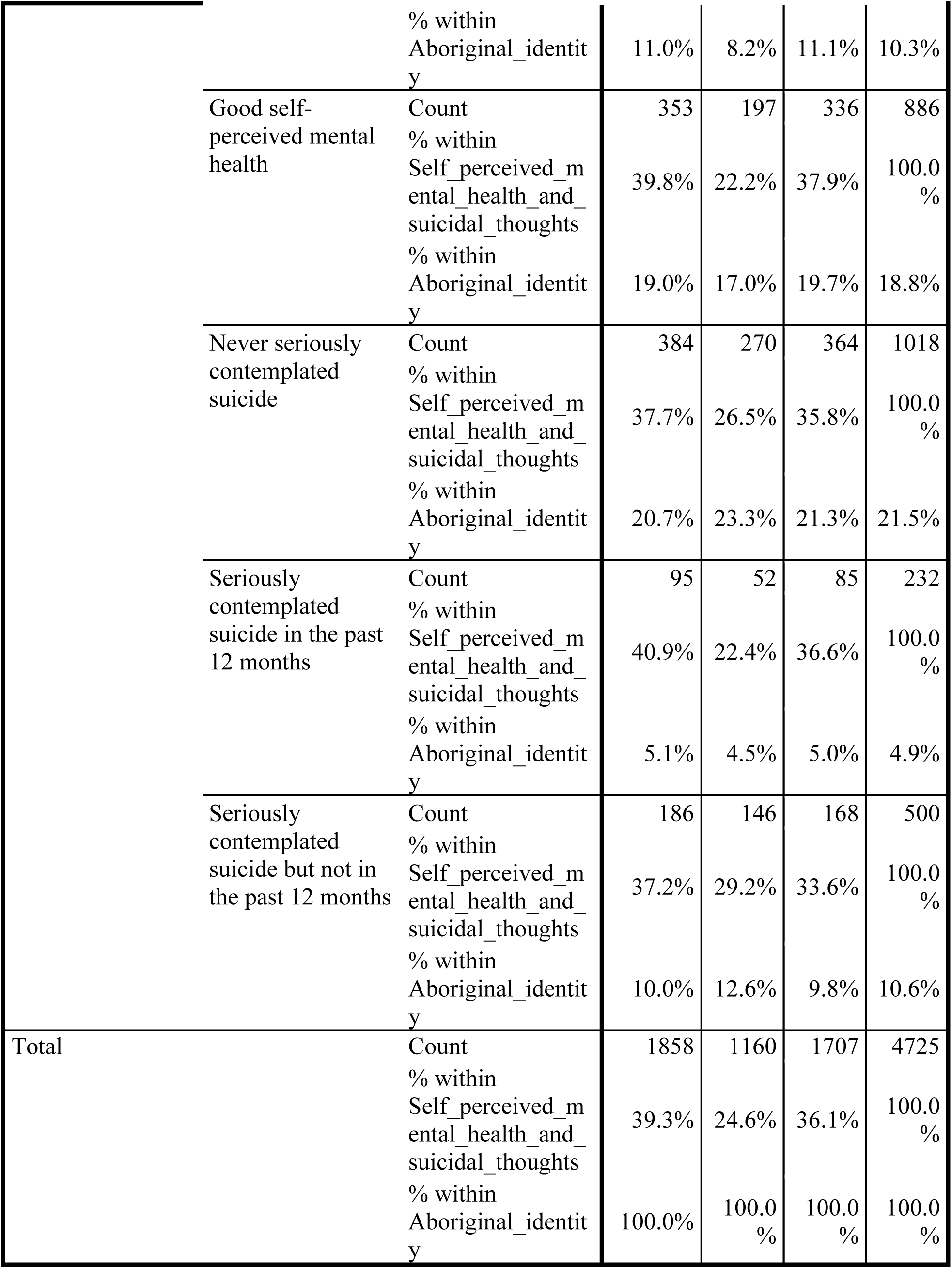
Crosstab.

**Crosstab**
|  |  |  | Aboriginal_identity |  |  | Total |
| --- | --- | --- | --- | --- | --- | --- |
|  |  |  | First nation | Inuit | Metis |  |
| Self_perceived_mental_health_and_suicidal_thoughts | Ever seriously contemplated suicide | Count | 251 | 172 | 209 | 632 |
|  |  | % within Self_perceived_mental_health_and_suicidal_thoughts | 39.7% | 27.2% | 33.1% | 100.0% |
|  |  | % within Aboriginal_identity | 13.5% | 14.8% | 12.2% | 13.4% |
|  | Excellent or very good self-perceived mental health | Count | 384 | 228 | 356 | 968 |
|  |  | % within Self_perceived_mental_health_and_suicidal_thoughts | 39.7% | 23.6% | 36.8% | 100.0% |
|  |  | % within Aboriginal_identity | 20.7% | 19.7% | 20.9% | 20.5% |
|  | Fair or poor self-perceived mental health | Count | 205 | 95 | 189 | 489 |
|  |  | % within Self_perceived_mental_health_and_suicidal_thoughts | 41.9% | 19.4% | 38.7% | 100.0% |

|  |  |  |  |  |  |
| --- | --- | --- | --- | --- | --- |
|  | % within<br>Aboriginal_identity | 11.0% | 8.2% | 11.1% | 10.3% |
| Good self-perceived mental health | Count<br>% within<br>Self_perceived_mental_health_and_suicidal_thoughts<br>% within<br>Aboriginal_identity | 353<br>39.8%<br>19.0% | 197<br>22.2%<br>17.0% | 336<br>37.9%<br>19.7% | 886<br>100.0%<br>18.8% |
| Never seriously contemplated suicide | Count<br>% within<br>Self_perceived_mental_health_and_suicidal_thoughts<br>% within<br>Aboriginal_identity | 384<br>37.7%<br>20.7% | 270<br>26.5%<br>23.3% | 364<br>35.8%<br>21.3% | 1018<br>100.0%<br>21.5% |
| Seriously contemplated suicide in the past 12 months | Count<br>% within<br>Self_perceived_mental_health_and_suicidal_thoughts<br>% within<br>Aboriginal_identity | 95<br>40.9%<br>5.1% | 52<br>22.4%<br>4.5% | 85<br>36.6%<br>5.0% | 232<br>100.0%<br>4.9% |
| Seriously contemplated suicide but not in the past 12 months | Count<br>% within<br>Self_perceived_mental_health_and_suicidal_thoughts<br>% within<br>Aboriginal_identity | 186<br>37.2%<br>10.0% | 146<br>29.2%<br>12.6% | 168<br>33.6%<br>9.8% | 500<br>100.0%<br>10.6% |
| Total | Count<br>% within<br>Self_perceived_mental_health_and_suicidal_thoughts<br>% within<br>Aboriginal_identity | 1858<br>39.3%<br>100.0% | 1160<br>24.6%<br>100.0% | 1707<br>36.1%<br>100.0% | 4725<br>100.0%<br>100.0% |

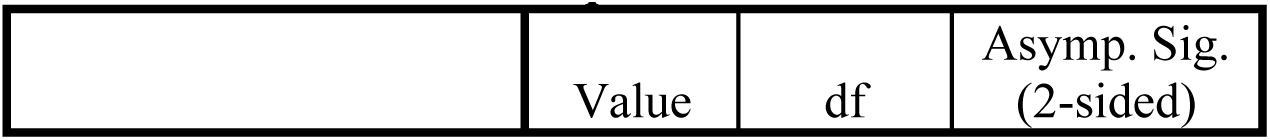
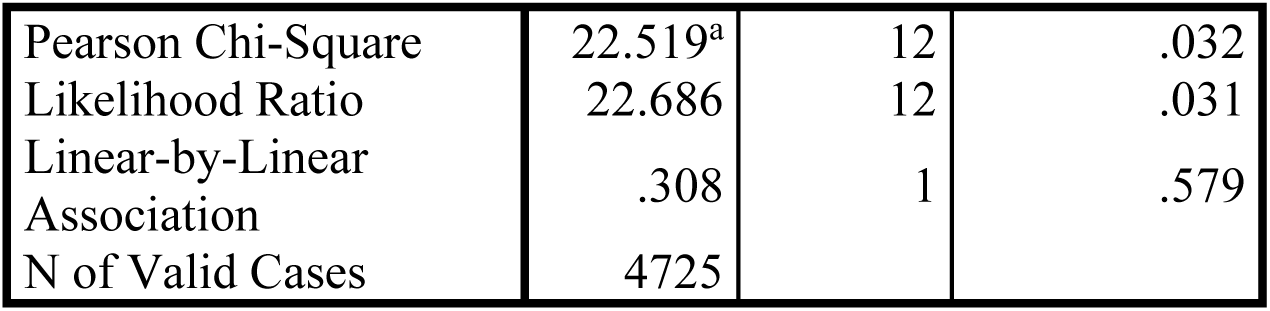
Chi-Square Tests.

Chi-Square Tests
|  | Value | df | Asymp. Sig.<br>(2-sided) |
| --- | --- | --- | --- |
| Pearson Chi-Square | 22.519<sup>a</sup> | 12 | .032 |
| Likelihood Ratio | 22.686 | 12 | .031 |
| Linear-by-Linear Association | .308 | 1 | .579 |
| N of Valid Cases | 4725 |  |  |

a. 0 cells (.0%) have expected count less than 5. The minimum expected count is 56.96.

Symmetric Measures.

Symmetric Measures
|  |  | Value | Asymp. Std. Error<sup>a</sup> | Approx. T<sup>b</sup> | Approx. Sig. |
| --- | --- | --- | --- | --- | --- |
| Interval by Interval | Pearson's R | .008 | .014 | .555 | .579<sup>c</sup> |
| Ordinal by Ordinal | Spearman Correlation | .009 | .014 | .619 | .536<sup>c</sup> |
| N of Valid Cases |  | 4725 |  |  |  |

a. Not assuming the null hypothesis.
b. Using the asymptotic standard error assuming the null hypothesis.
c. Based on normal approximation.

a. Not assuming the null hypothesis.
b. Using the asymptotic standard error assuming the null hypothesis.
c. Based on normal approximation.

**Self_perceived_mental_health_and_suicidal_thoughts * Age_group**

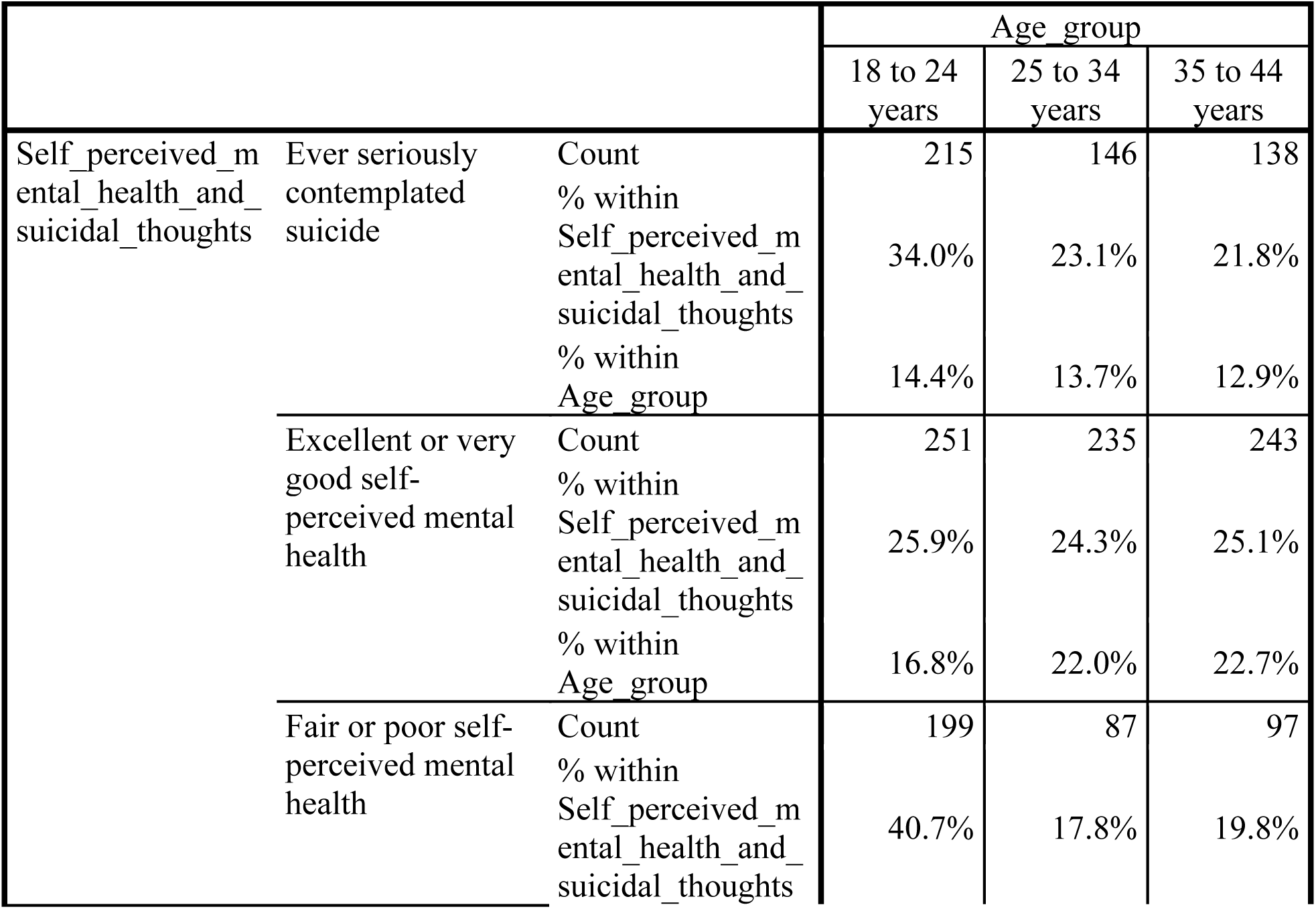
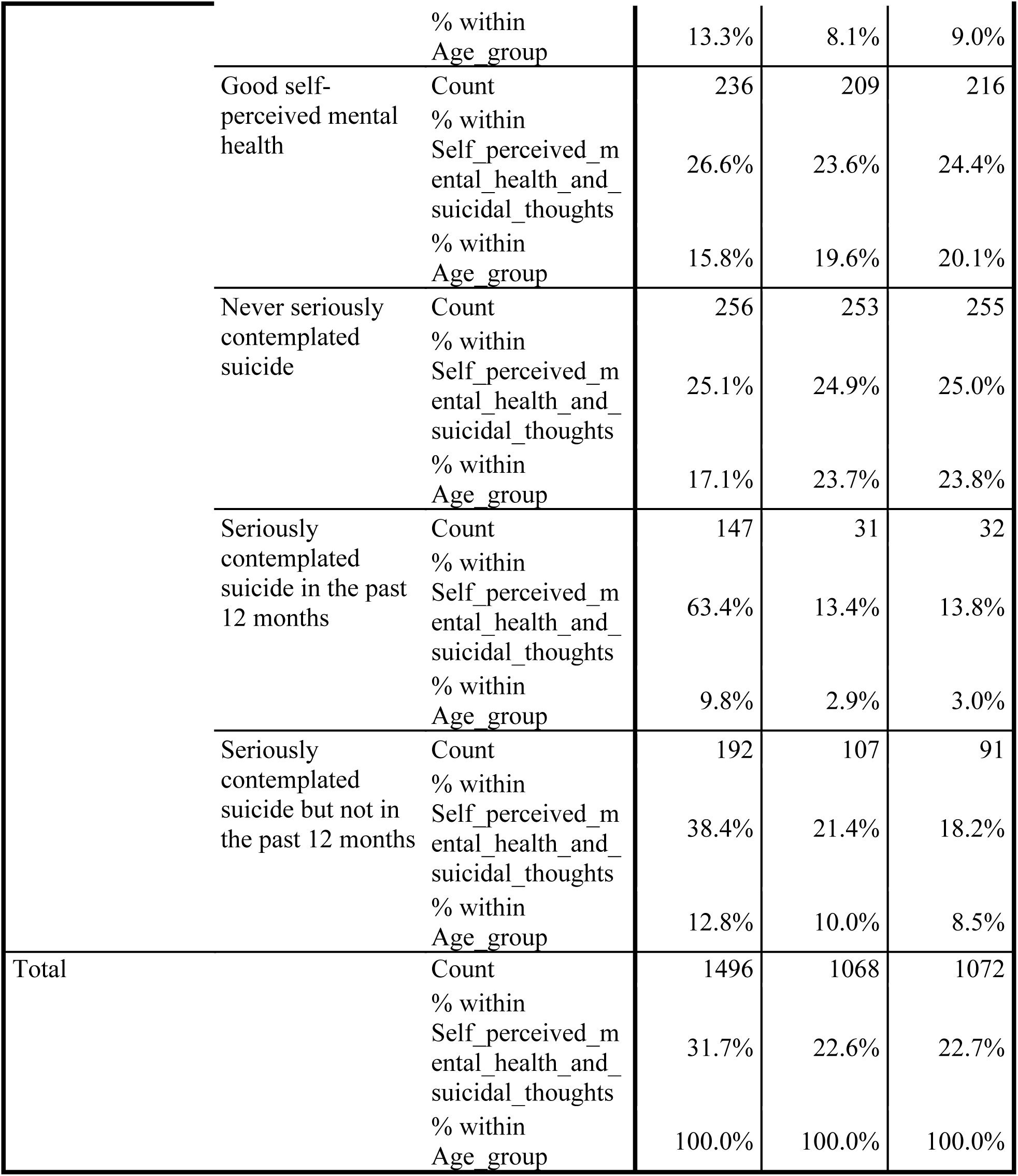
Crosstab.

Self\_perceived\_mental\_health\_and\_suicidal\_thoughts \* Age\_group Crosstab
|  |  |  | Age_group |  |  |
| --- | --- | --- | --- | --- | --- |
|  |  |  | 18 to 24 years | 25 to 34 years | 35 to 44 years |
| Self_perceived_mental_health_and_suicidal_thoughts | Ever seriously contemplated suicide | Count | 215 | 146 | 138 |
|  |  | % within Self_perceived_mental_health_and_suicidal_thoughts | 34.0% | 23.1% | 21.8% |
|  |  | % within Age_group | 14.4% | 13.7% | 12.9% |
|  | Excellent or very good self-perceived mental health | Count | 251 | 235 | 243 |
|  |  | % within Self_perceived_mental_health_and_suicidal_thoughts | 25.9% | 24.3% | 25.1% |
|  |  | % within Age_group | 16.8% | 22.0% | 22.7% |
|  | Fair or poor self-perceived mental health | Count | 199 | 87 | 97 |
|  |  | % within Self_perceived_mental_health_and_suicidal_thoughts | 40.7% | 17.8% | 19.8% |

|  |  |  |  |  |
| --- | --- | --- | --- | --- |
|  | % within<br>Age_group | 13.3% | 8.1% | 9.0% |
| Good self-perceived mental health | Count | 236 | 209 | 216 |
|  | % within<br>Self_perceived_mental_health_and_suicidal_thoughts | 26.6% | 23.6% | 24.4% |
|  | % within<br>Age_group | 15.8% | 19.6% | 20.1% |
| Never seriously contemplated suicide | Count | 256 | 253 | 255 |
|  | % within<br>Self_perceived_mental_health_and_suicidal_thoughts | 25.1% | 24.9% | 25.0% |
|  | % within<br>Age_group | 17.1% | 23.7% | 23.8% |
| Seriously contemplated suicide in the past 12 months | Count | 147 | 31 | 32 |
|  | % within<br>Self_perceived_mental_health_and_suicidal_thoughts | 63.4% | 13.4% | 13.8% |
|  | % within<br>Age_group | 9.8% | 2.9% | 3.0% |
| Seriously contemplated suicide but not in the past 12 months | Count | 192 | 107 | 91 |
|  | % within<br>Self_perceived_mental_health_and_suicidal_thoughts | 38.4% | 21.4% | 18.2% |
|  | % within<br>Age_group | 12.8% | 10.0% | 8.5% |
| Total | Count | 1496 | 1068 | 1072 |
|  | % within<br>Self_perceived_mental_health_and_suicidal_thoughts | 31.7% | 22.6% | 22.7% |
|  | % within<br>Age_group | 100.0% | 100.0% | 100.0% |

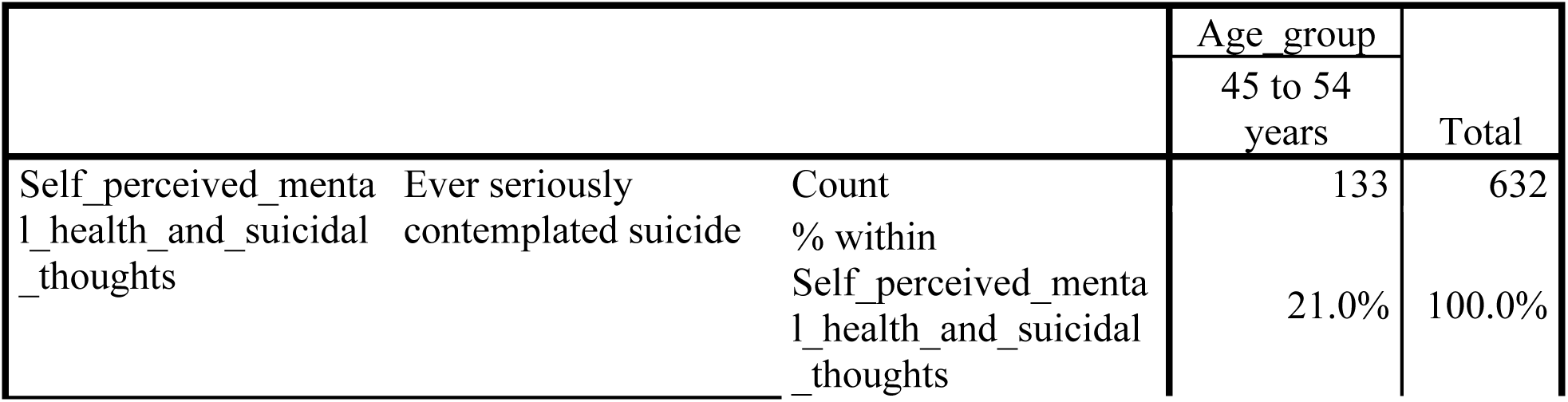
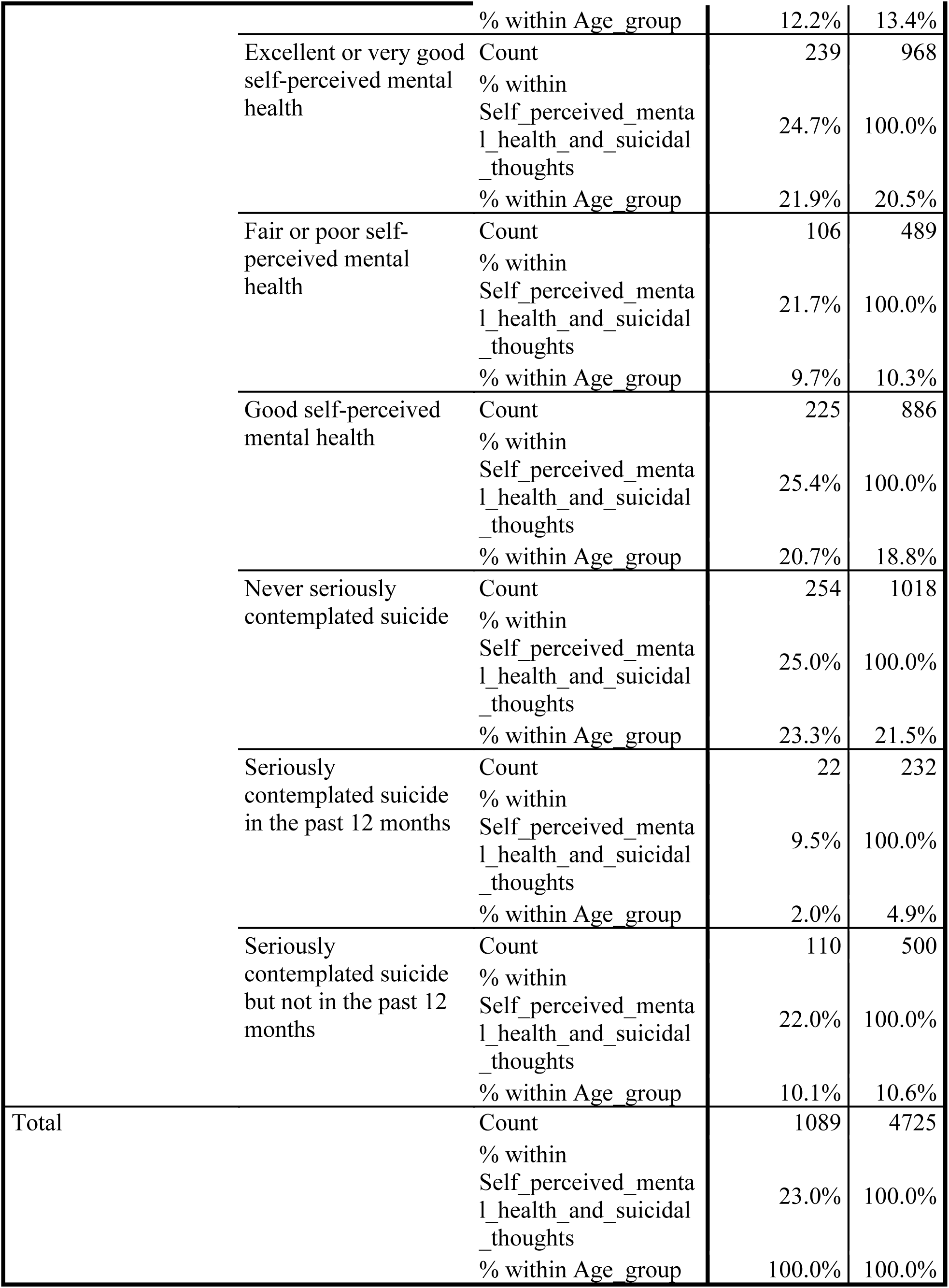
Crosstab.

Crosstab
|  |  |  | Age_group |  |
| --- | --- | --- | --- | --- |
|  |  |  | 45 to 54 years | Total |
| Self_perceived_mental_health_and_suicidal_thoughts | Ever seriously contemplated suicide | Count | 133 | 632 |
|  |  | % within<br>Self_perceived_mental_health_and_suicidal_thoughts | 21.0% | 100.0% |

|  |  |  |  |
| --- | --- | --- | --- |
|  | % within Age_group | 12.2% | 13.4% |
| Excellent or very good self-perceived mental health | Count | 239 | 968 |
|  | % within Self_perceived_mental_health_and_suicidal_thoughts | 24.7% | 100.0% |
|  | % within Age_group | 21.9% | 20.5% |
| Fair or poor self-perceived mental health | Count | 106 | 489 |
|  | % within Self_perceived_mental_health_and_suicidal_thoughts | 21.7% | 100.0% |
|  | % within Age_group | 9.7% | 10.3% |
| Good self-perceived mental health | Count | 225 | 886 |
|  | % within Self_perceived_mental_health_and_suicidal_thoughts | 25.4% | 100.0% |
|  | % within Age_group | 20.7% | 18.8% |
| Never seriously contemplated suicide | Count | 254 | 1018 |
|  | % within Self_perceived_mental_health_and_suicidal_thoughts | 25.0% | 100.0% |
|  | % within Age_group | 23.3% | 21.5% |
| Seriously contemplated suicide in the past 12 months | Count | 22 | 232 |
|  | % within Self_perceived_mental_health_and_suicidal_thoughts | 9.5% | 100.0% |
|  | % within Age_group | 2.0% | 4.9% |
| Seriously contemplated suicide but not in the past 12 months | Count | 110 | 500 |
|  | % within Self_perceived_mental_health_and_suicidal_thoughts | 22.0% | 100.0% |
|  | % within Age_group | 10.1% | 10.6% |
| Total | Count | 1089 | 4725 |
|  | % within Self_perceived_mental_health_and_suicidal_thoughts | 23.0% | 100.0% |
|  | % within Age_group | 100.0% | 100.0% |

Chi-Square Tests.

Chi-Square Tests
|  | Value | df | Asymp. Sig. (2-sided) |
| --- | --- | --- | --- |
| Pearson Chi-Square | 189.001<sup>a</sup> | 18 | .000 |
| Likelihood Ratio | 181.437 | 18 | .000 |
| Linear-by-Linear Association | 7.400 | 1 | .007 |
| N of Valid Cases | 4725 |  |  |

a. 0 cells (.0%) have expected count less than 5. The minimum expected count is 52.44.

Symmetric Measures.

Symmetric Measures
|  |  | Value | Asymp. Std. Error<sup>a</sup> | Approx. T<sup>b</sup> | Approx. Sig. |
| --- | --- | --- | --- | --- | --- |
| Interval by Interval | Pearson's R | -.040 | .015 | -2.722 | .007<sup>c</sup> |
| Ordinal by Ordinal | Spearman Correlation | -.037 | .015 | -2.530 | .011<sup>c</sup> |
| N of Valid Cases |  | 4725 |  |  |  |

a. Not assuming the null hypothesis.
b. Using the asymptotic standard error assuming the null hypothesis.
c. Based on normal approximation.

a. Not assuming the null hypothesis.
b. Using the asymptotic standard error assuming the null hypothesis.
c. Based on normal approximation.

**Self_perceived_mental_health_and_suicidal_thoughts * Sex**

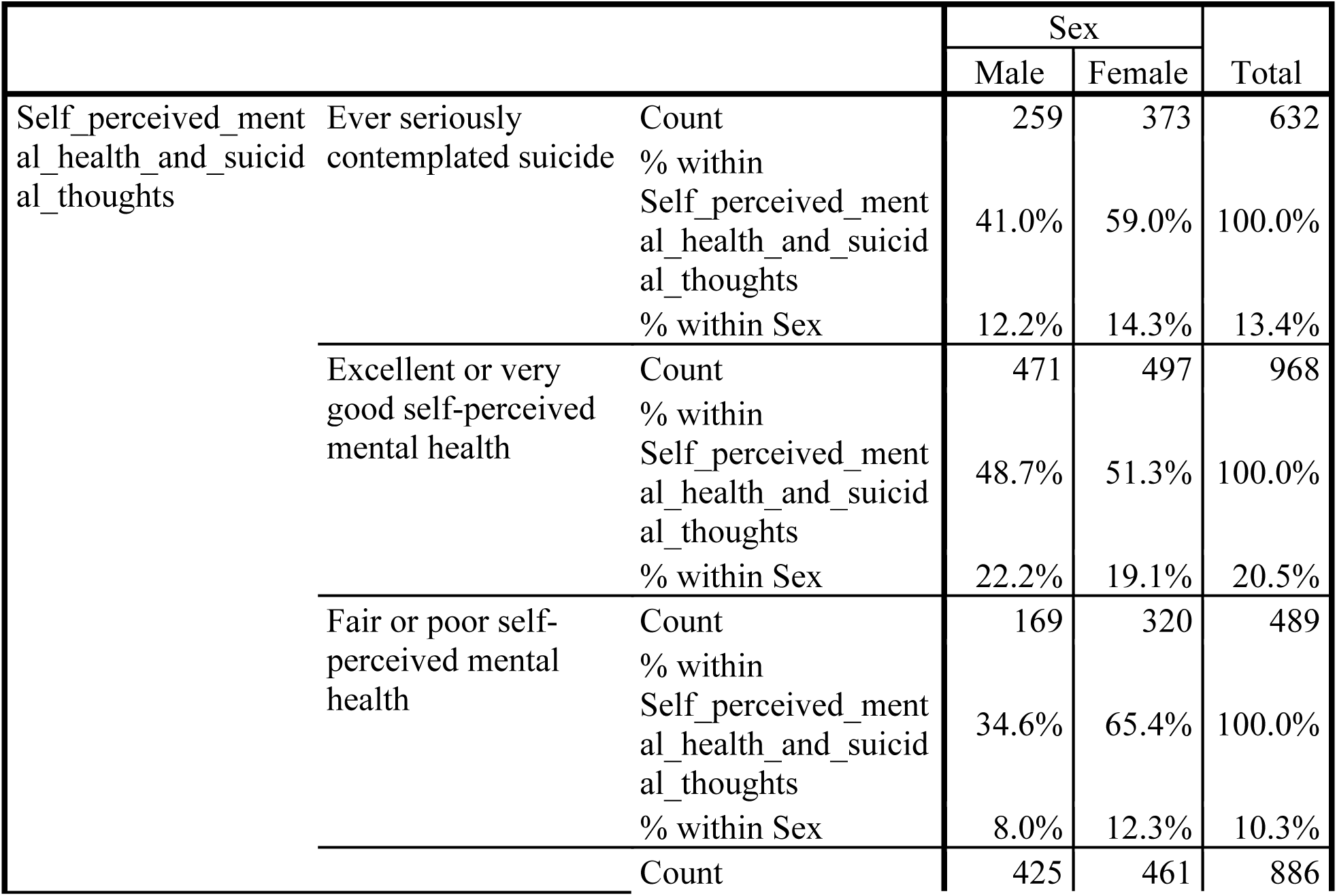
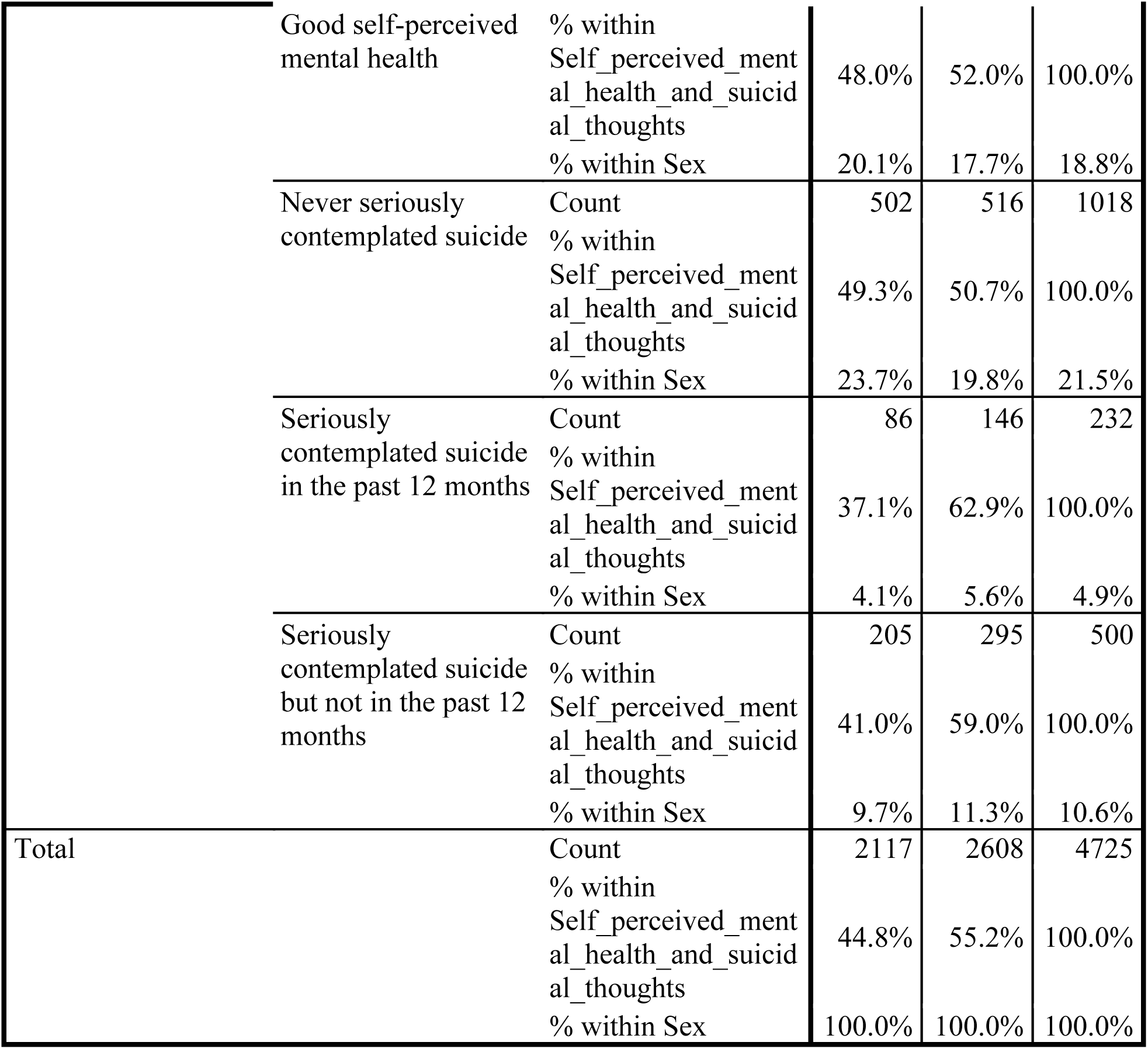
Crosstab.

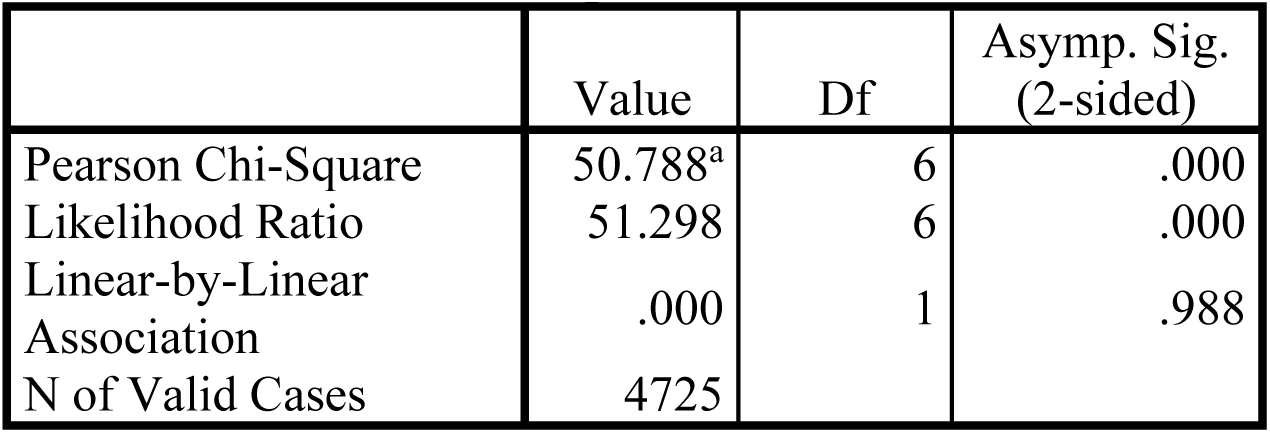
Chi-Square Tests.

Chi-Square Tests
|  | Value | Df | Asymp. Sig. (2-sided) |
| --- | --- | --- | --- |
| Pearson Chi-Square | 50.788<sup>a</sup> | 6 | .000 |
| Likelihood Ratio | 51.298 | 6 | .000 |
| Linear-by-Linear Association | .000 | 1 | .988 |
| N of Valid Cases | 4725 |  |  |

a. 0 cells (.0%) have expected count less than 5. The minimum expected count is 103.95.

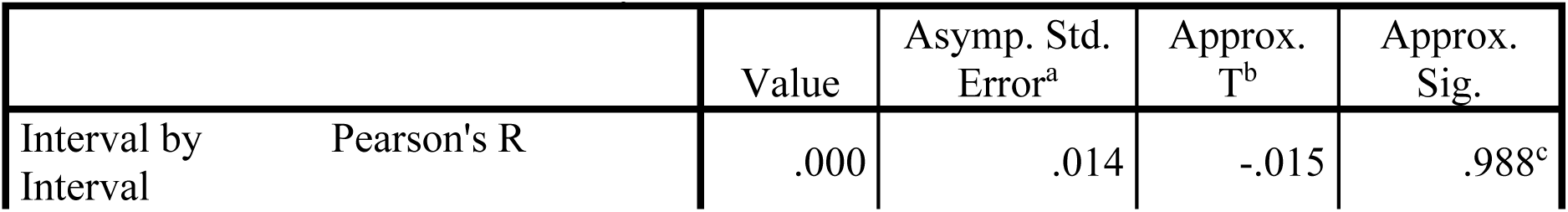
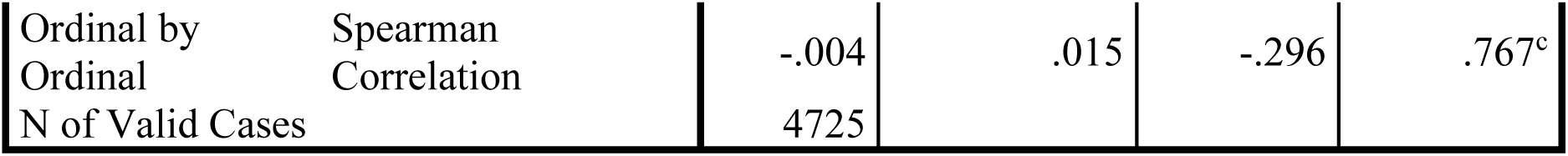
Symmetric Measures.

a. Not assuming the null hypothesis.
b. Using the asymptotic standard error assuming the null hypothesis.
c. Based on normal approximation.

CROSSTABS

/TABLES=Aboriginal_identity Age_group Sex BY Suicidal_Thoughts

/FORMAT=AVALUE TABLES

/STATISTICS=CHISQ CORR

/CELLS=COUNT ROW COLUMN

/COUNT ROUND CELL.

**Crosstabs**

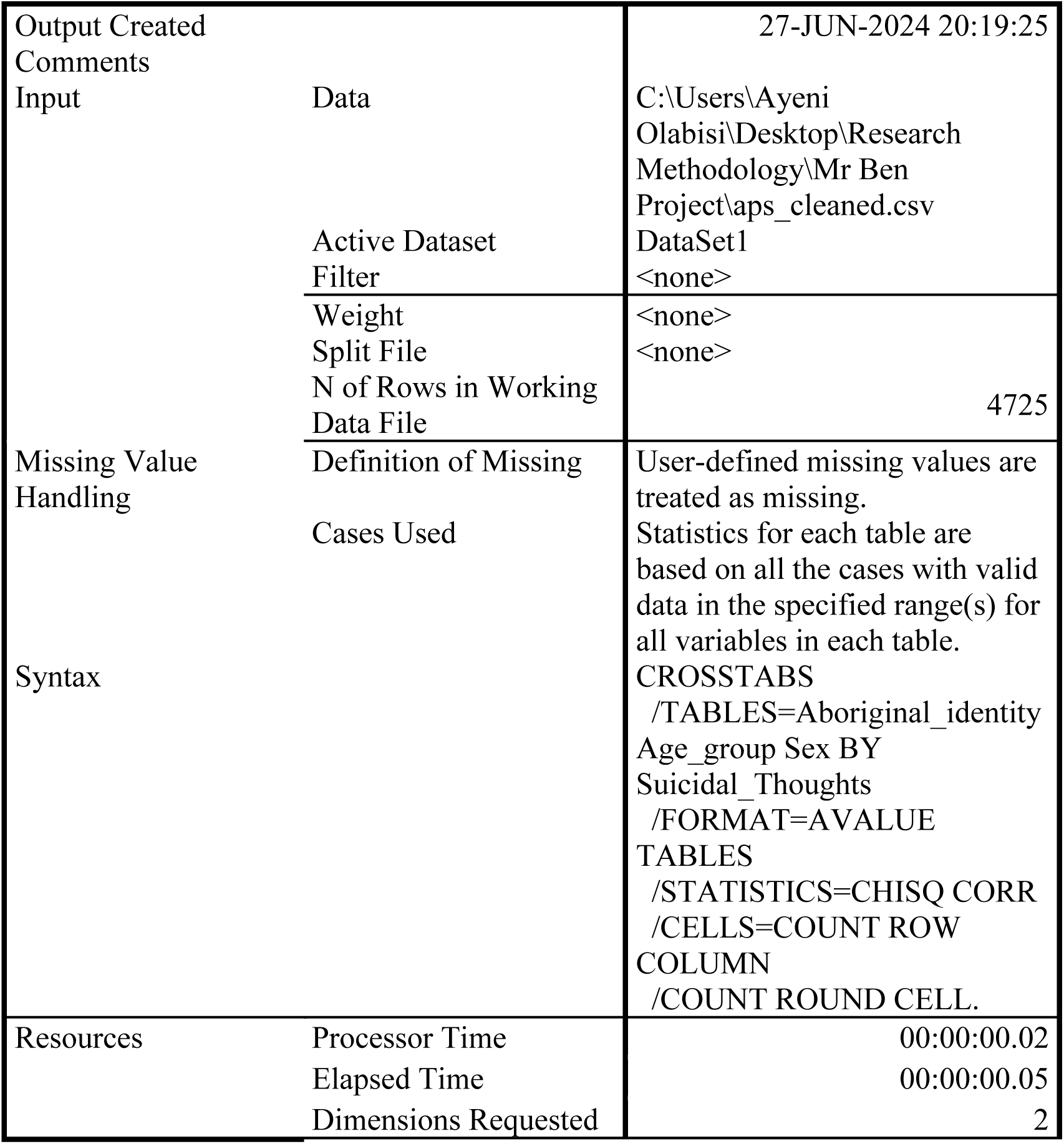
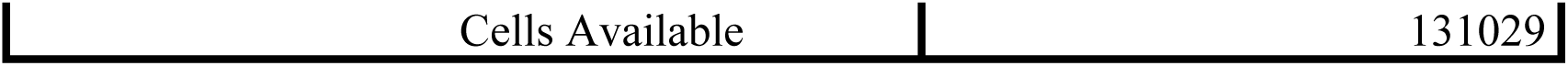
Notes.

Notes
|  |  |  |
| --- | --- | --- |
| Output Created |  | 27-JUN-2024 20:19:25 |
| Comments |  |  |
| Input | Data | C:\Users\Ayeni<br>Olabisi\Desktop\Research<br>Methodology\Mr Ben<br>Project\aps_cleaned.csv |
|  | Active Dataset | DataSet1 |
|  | Filter | <none> |
|  | Weight | <none> |
|  | Split File | <none> |
|  | N of Rows in Working<br>Data File | 4725 |
| Missing Value<br>Handling | Definition of Missing | User-defined missing values are<br>treated as missing. |
|  | Cases Used | Statistics for each table are<br>based on all the cases with valid<br>data in the specified range(s) for<br>all variables in each table. |
| Syntax |  | CROSSTABS<br>/TABLES=Aboriginal_identity<br>Age_group Sex BY<br>Suicidal_Thoughts<br>/FORMAT=AVALUE<br>TABLES<br>/STATISTICS=CHISQ CORR<br>/CELLS=COUNT ROW<br>COLUMN<br>/COUNT ROUND CELL. |
| Resources | Processor Time | 00:00:00.02 |
|  | Elapsed Time | 00:00:00.05 |
|  | Dimensions Requested | 2 |

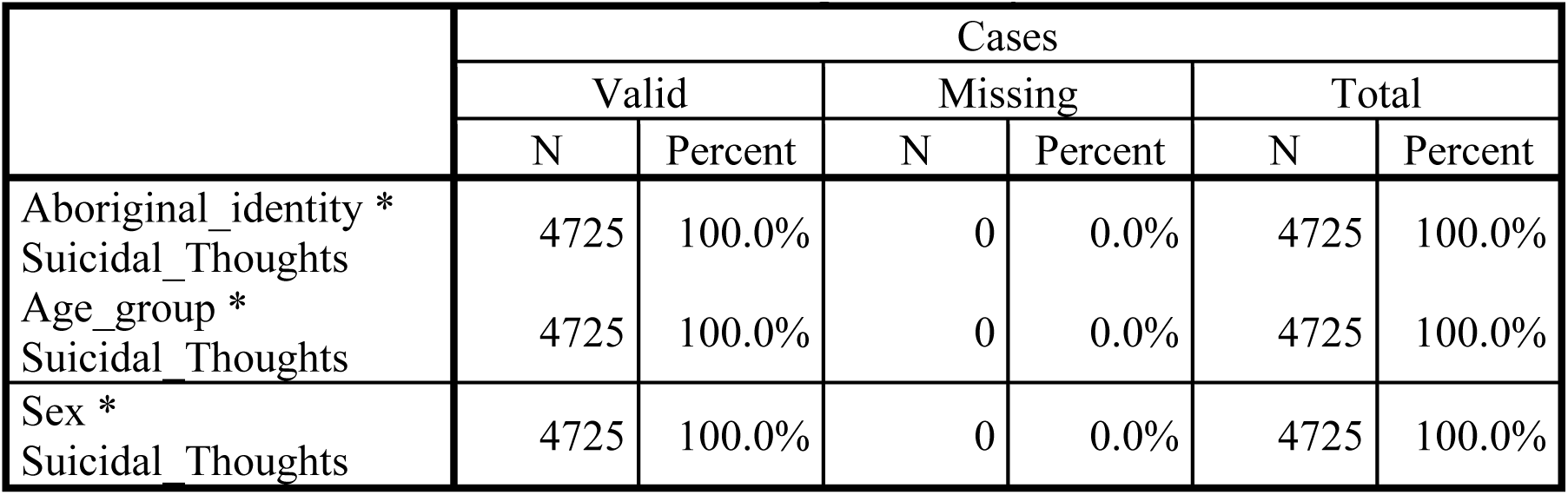
Case Processing Summary.

**Aboriginal_identity * Suicidal_Thoughts**

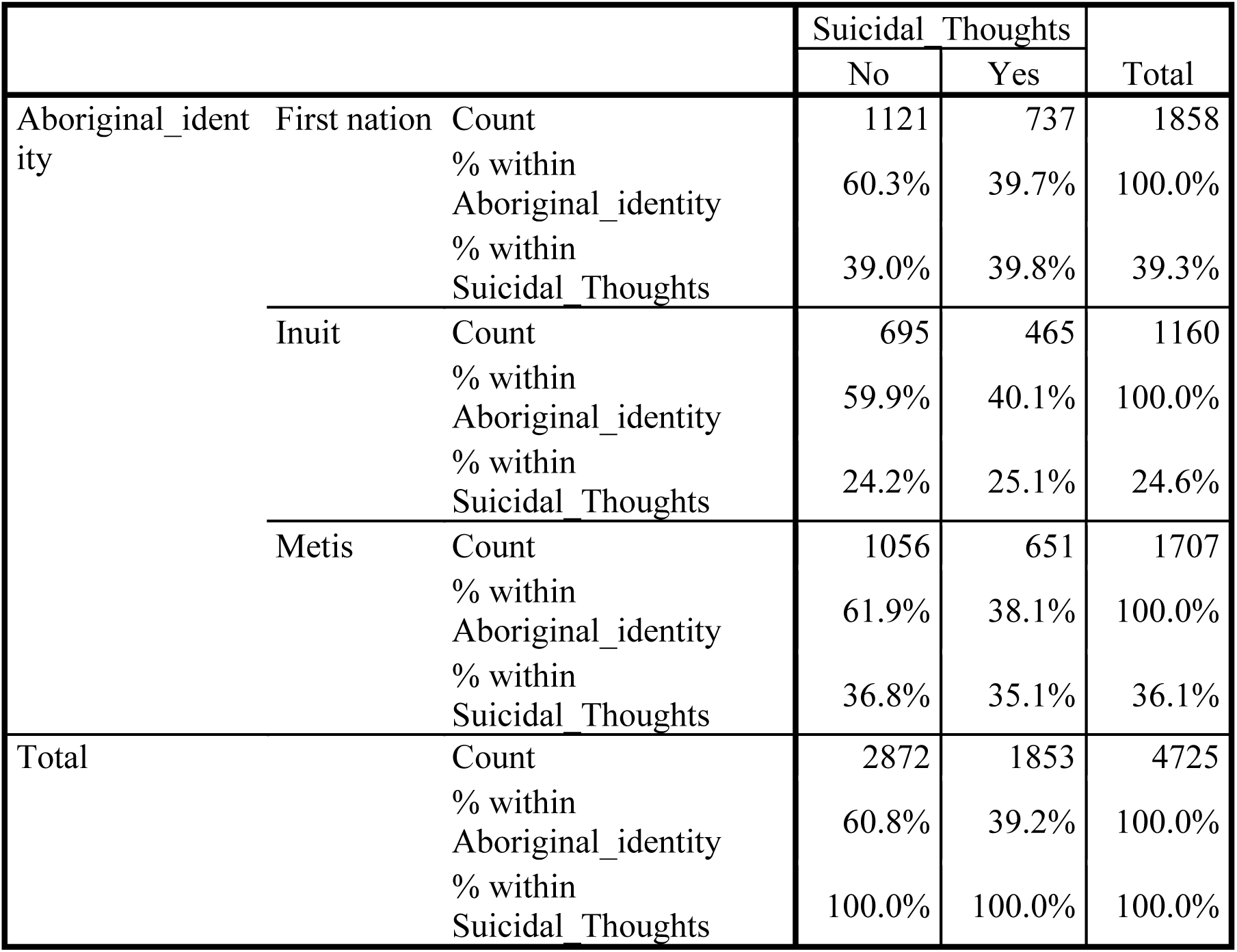
Crosstab.

Chi-Square Tests.

**Chi-Square Tests**
|  | Value | df | Asymp. Sig. (2-sided) |
| --- | --- | --- | --- |
| Pearson Chi-Square | 1.360<sup>a</sup> | 2 | .507 |
| Likelihood Ratio | 1.362 | 2 | .506 |
| Linear-by-Linear Association | .845 | 1 | .358 |
| N of Valid Cases | 4725 |  |  |

a. 0 cells (.0%) have expected count less than 5. The minimum expected count is 454.92.

Symmetric Measures.

Symmetric Measures
|  |  | Value | Asymp. Std. Error<sup>a</sup> | Approx. T<sup>b</sup> | Approx. Sig. |
| --- | --- | --- | --- | --- | --- |
| Interval by Interval | Pearson's R | -.013 | .015 | -.919 | .358<sup>c</sup> |
| Ordinal by Ordinal | Spearman Correlation | -.013 | .015 | -.910 | .363<sup>c</sup> |
| N of Valid Cases |  | 4725 |  |  |  |

a. Not assuming the null hypothesis.
b. Using the asymptotic standard error assuming the null hypothesis.
c. Based on normal approximation.

a. Not assuming the null hypothesis.
b. Using the asymptotic standard error assuming the null hypothesis.
c. Based on normal approximation.

**Age_group * Suicidal_Thoughts**

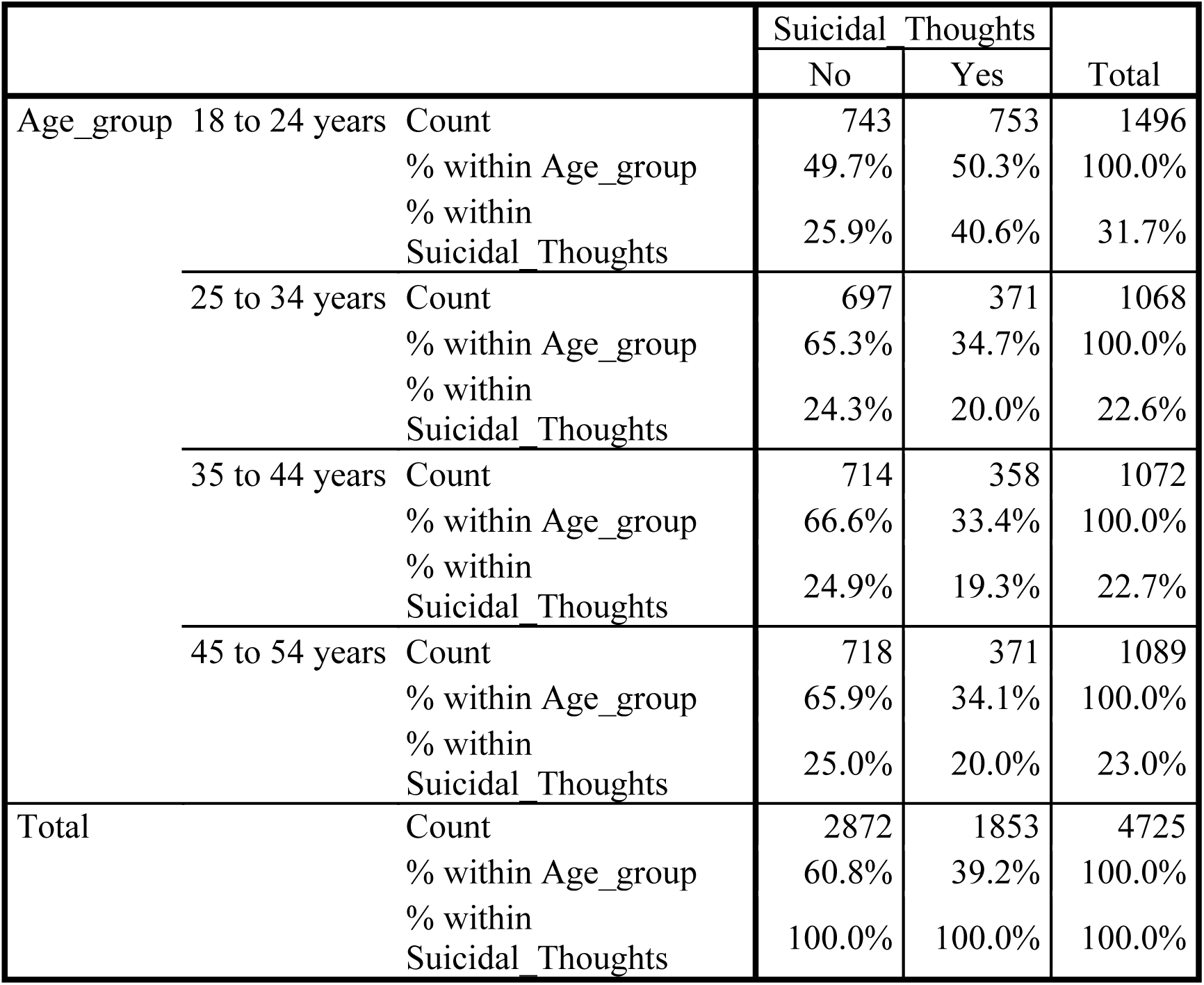
Crosstab.

Chi-Square Tests.

Chi-Square Tests
|  | Value | df | Asymp. Sig. (2-sided) |
| --- | --- | --- | --- |
| Pearson Chi-Square | 113.907<sup>a</sup> | 3 | .000 |
| Likelihood Ratio | 112.706 | 3 | .000 |
| Linear-by-Linear Association | 77.687 | 1 | .000 |
| N of Valid Cases | 4725 |  |  |

a. 0 cells (.0%) have expected count less than 5. The minimum expected count is 418.84.

Symmetric Measures.

Symmetric Measures
|  |  | Value | Asymp. Std. Error<sup>a</sup> | Approx. T<sup>b</sup> | Approx. Sig. |
| --- | --- | --- | --- | --- | --- |
| Interval by Interval | Pearson's R | -.128 | .015 | -8.886 | .000<sup>c</sup> |
| Ordinal by Ordinal | Spearman Correlation | -.132 | .015 | -9.138 | .000<sup>c</sup> |
| N of Valid Cases |  | 4725 |  |  |  |

a. Not assuming the null hypothesis.
b. Using the asymptotic standard error assuming the null hypothesis.
c. Based on normal approximation.

a. Not assuming the null hypothesis.
b. Using the asymptotic standard error assuming the null hypothesis.
c. Based on normal approximation.

**Sex * Suicidal_Thoughts**

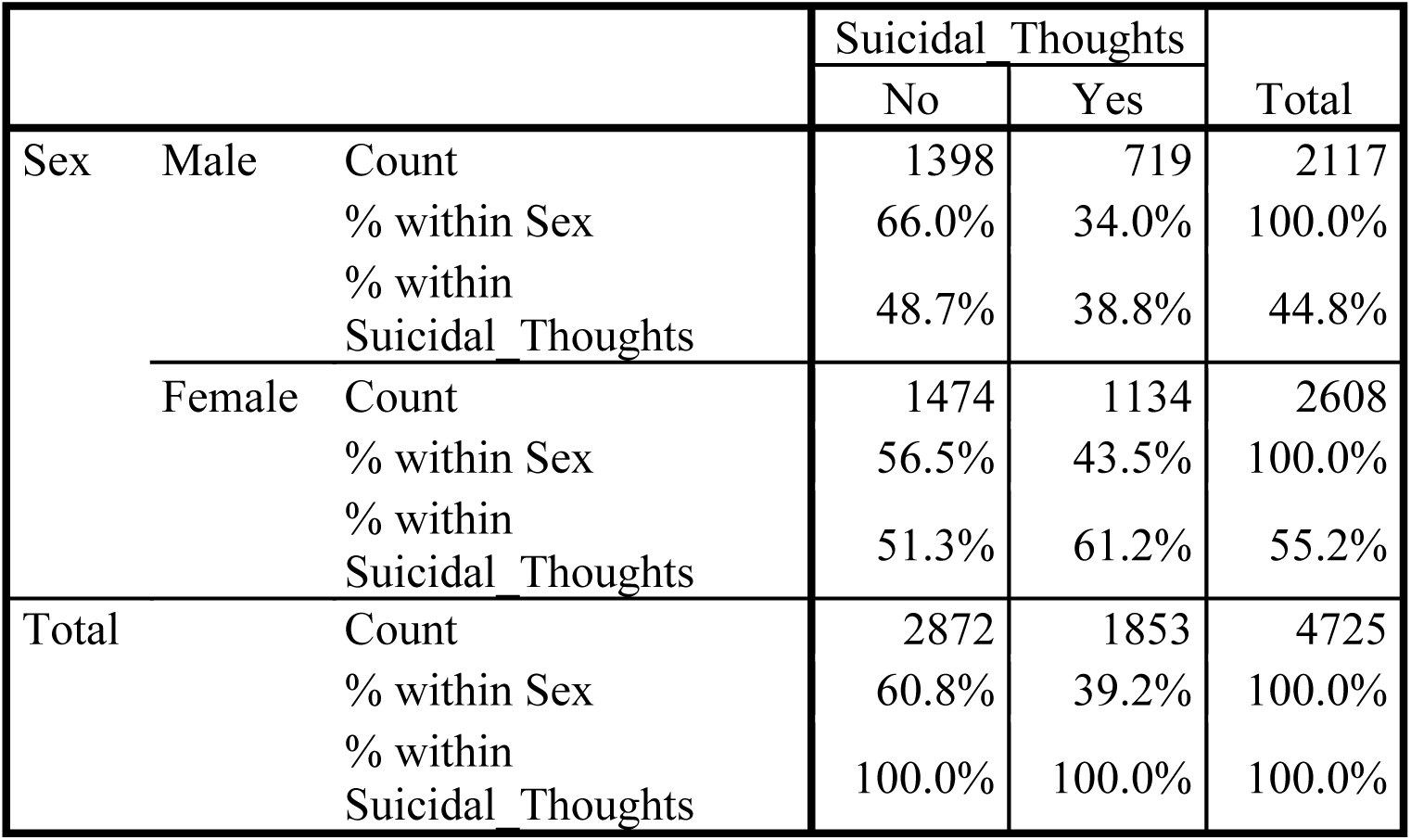
Crosstab.

Chi-Square Tests.

Chi-Square Tests
|  | Value | df | Asymp. Sig. (2-sided) | Exact Sig. (2-sided) | Exact Sig. (1-sided) |
| --- | --- | --- | --- | --- | --- |
| Pearson Chi-Square | 44.412<sup>a</sup> | 1 | .000 |  |  |
| Continuity Correction<sup>b</sup> | 44.014 | 1 | .000 |  |  |
| Likelihood Ratio | 44.634 | 1 | .000 |  |  |
| Fisher's Exact Test |  |  |  | .000 | .000 |
| Linear-by-Linear Association | 44.403 | 1 | .000 |  |  |
| N of Valid Cases | 4725 |  |  |  |  |

a. 0 cells (.0%) have expected count less than 5. The minimum expected count is 830.22.
b. Computed only for a 2×2 table

Symmetric Measures.

Symmetric Measures
|  |  | Value | Asymp. Std. Error<sup>a</sup> | Approx. T<sup>b</sup> | Approx. Sig. |
| --- | --- | --- | --- | --- | --- |
| Interval by Interval | Pearson's R | .097 | .014 | 6.694 | .000<sup>c</sup> |
| Ordinal by Ordinal | Spearman Correlation | .097 | .014 | 6.694 | .000<sup>c</sup> |
| N of Valid Cases |  | 4725 |  |  |  |

a. Not assuming the null hypothesis.
b. Using the asymptotic standard error assuming the null hypothesis.
c. Based on normal approximation.

LOGISTIC REGRESSION VARIABLES Suicidal_Thoughts

/METHOD=ENTER Self_perceived_mental_health_and_suicidal_thoughts

Aboriginal_identity Age_group Sex SES

/CONTRAST (Aboriginal_identity)=Indicator

/PRINT=GOODFIT CI(95)

/CRITERIA=PIN(0.05) POUT(0.10) ITERATE(20) CUT(0.5).

**Logistic Regression**

Case Processing Summary.

Case Processing Summary
| Unweighted Cases<sup>a</sup> | N | Percent |
| --- | --- | --- |
| Selected Cases |  |  |
| Included in Analysis | 4725 | 100.0 |
| Missing Cases | 0 | .0 |
| Total | 4725 | 100.0 |
| Unselected Cases | 0 | .0 |
| Total | 4725 | 100.0 |

a. If weight is in effect, see classification table for the total number of cases.

**Dependent Variable**

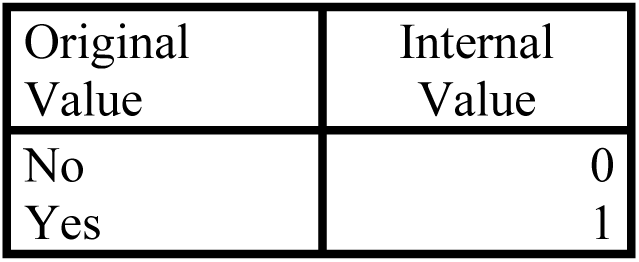
Encoding.

Dependent Variable Encoding
| Original Value | Internal Value |
| --- | --- |
| No | 0 |
| Yes | 1 |

Categorical Variables Codings.

Categorical Variables Codings
|  |  | Frequency | Parameter coding |  |
| --- | --- | --- | --- | --- |
|  |  |  | (1) | (2) |
| Aboriginal_identity | First nation | 1858 | 1.000 | .000 |
|  | Inuit | 1160 | .000 | 1.000 |
|  | Metis | 1707 | .000 | .000 |

**Block 0: Beginning Block**

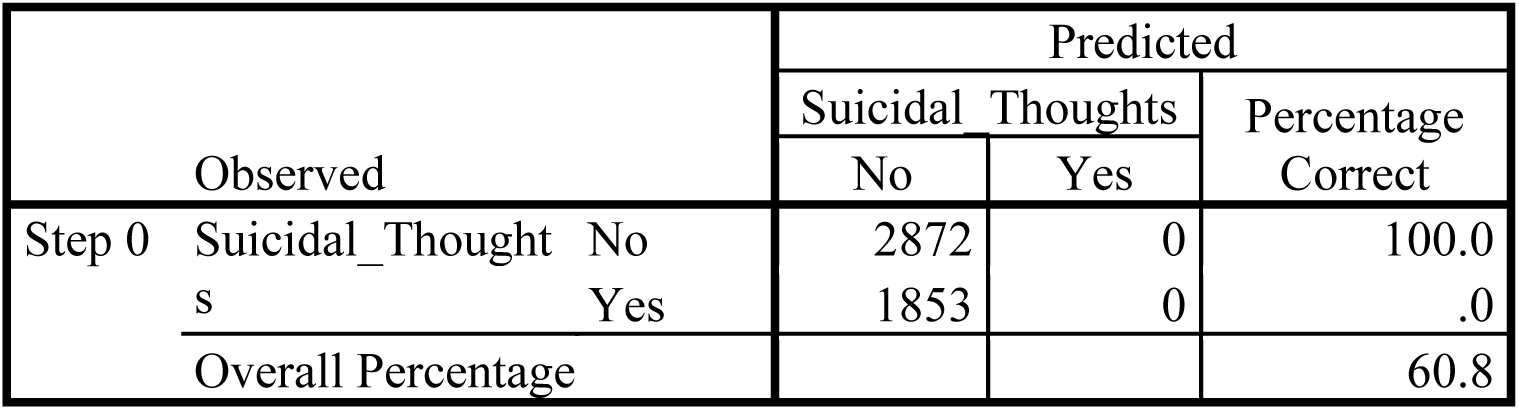
Classification Table^a,b^.

a. Constant is included in the model.
b. The cut value is .500

Variables in the Equation.

Variables in the Equation
|  | B | S.E. | Wald | df | Sig. | Exp(B) |
| --- | --- | --- | --- | --- | --- | --- |
| Step 0 Constant | -.438 | .030 | 216.276 | 1 | .000 | .645 |

Variables not in the Equation.

Variables not in the Equation
|  |  |  | Score | df | Sig. |
| --- | --- | --- | --- | --- | --- |
| Step 0 | Variables | Self_perceived_mental_health_and_suicidal_thoughts | 2.822 | 1 | .093 |
|  |  | Aboriginal_identity | 1.360 | 2 | .507 |
|  |  | Aboriginal_identity(1) | .259 | 1 | .611 |
|  |  | Aboriginal_identity(2) | .487 | 1 | .485 |
|  |  | Age_group | 77.704 | 1 | .000 |
|  |  | Sex | 44.412 | 1 | .000 |
|  |  | SES | 38.549 | 1 | .000 |
|  | Overall Statistics |  | 161.746 | 6 | .000 |

**Block 1: Method = Enter**

Omnibus Tests of Model Coefficients.

Block 1: Method = Enter Omnibus Tests of Model Coefficients
|  |  | Chi-square | df | Sig. |
| --- | --- | --- | --- | --- |
| Step 1 | Step | 174.777 | 6 | .000 |
|  | Block | 174.777 | 6 | .000 |
|  | Model | 174.777 | 6 | .000 |

Model Summary.

Model Summary
| Step | -2 Log likelihood | Cox & Snell R Square | Nagelkerke R Square |
| --- | --- | --- | --- |
| 1 | 6153.969<sup>a</sup> | .036 | .049 |

a. Estimation terminated at iteration number 5 because parameter estimates changed by less than .001.

Hosmer and Lemeshow Test.

Hosmer and Lemeshow Test
| Step | Chi-square | df | Sig. |
| --- | --- | --- | --- |
| 1 | 65.715 | 8 | .000 |

Contingency Table for Hosmer and Lemeshow Test.

Contingency Table for Hosmer and Lemeshow Test
|  |  | Suicidal_Thoughts = No |  | Suicidal_Thoughts = Yes |  | Total |
| --- | --- | --- | --- | --- | --- | --- |
|  |  | Observed | Expected | Observed | Expected |  |
| Step 1 | 1 | 361 | 365.602 | 112 | 107.398 | 473 |
|  | 2 | 320 | 334.089 | 153 | 138.911 | 473 |
|  | 3 | 293 | 315.235 | 180 | 157.765 | 473 |
|  | 4 | 369 | 302.442 | 104 | 170.558 | 473 |
|  | 5 | 267 | 292.186 | 206 | 180.814 | 473 |
|  | 6 | 281 | 279.155 | 192 | 193.845 | 473 |
|  | 7 | 283 | 271.826 | 190 | 201.174 | 473 |
|  | 8 | 250 | 257.546 | 223 | 215.454 | 473 |
|  | 9 | 209 | 237.023 | 264 | 235.977 | 473 |
|  | 10 | 239 | 216.896 | 229 | 251.104 | 468 |

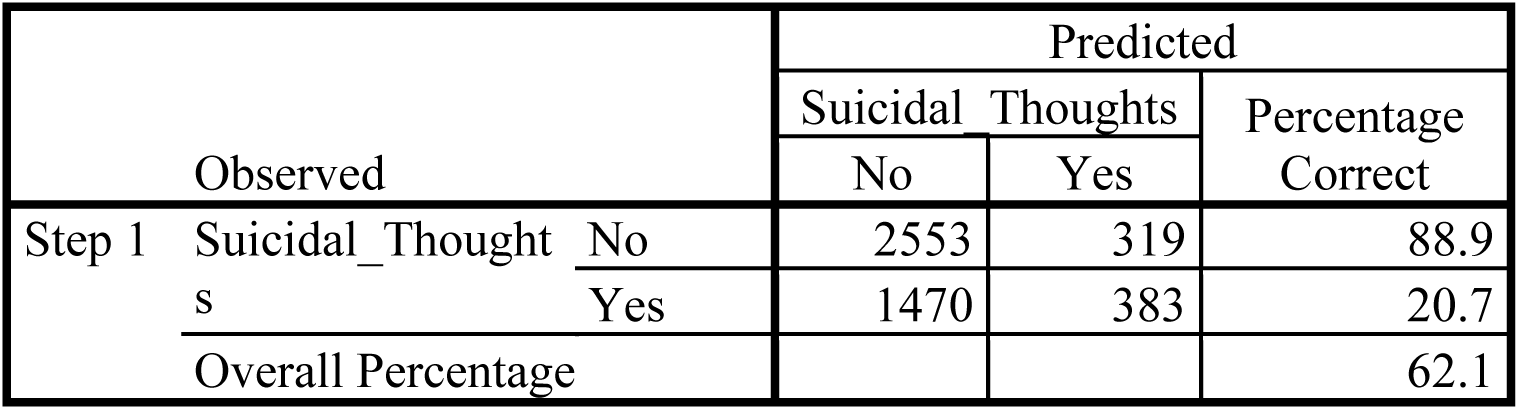
Classification Table^a^.

Classification Table<sup>a</sup>
| Observed |  |  | Predicted |  |  |
| --- | --- | --- | --- | --- | --- |
|  |  |  | Suicidal_Thoughts |  | Percentage Correct |
|  |  |  | No | Yes |  |
| Step 1 | Suicidal_Thoughts | No | 2553 | 319 | 88.9 |
|  |  | Yes | 1470 | 383 | 20.7 |
| Overall Percentage |  |  |  |  | 62.1 |

a. The cut value is .500

Variables in the Equation.

Variables in the Equation
|  | B | S.E. | Wald | df | Sig. | Exp(B) |
| --- | --- | --- | --- | --- | --- | --- |
| Step 1<sup>a</sup> Self_perceived_mental_health_and_suicidal_thoughts | .022 | .016 | 1.807 | 1 | .179 | 1.022 |
| Aboriginal_identity |  |  | .992 | 2 | .609 |  |
| Aboriginal_identity(1) | .065 | .070 | .846 | 1 | .358 | 1.067 |
| Aboriginal_identity(2) | .007 | .080 | .007 | 1 | .931 | 1.007 |
| Age_group | -.231 | .027 | 75.281 | 1 | .000 | .794 |
| Sex | .425 | .061 | 47.896 | 1 | .000 | 1.530 |
| SES | -.398 | .071 | 31.159 | 1 | .000 | .672 |
| Constant | -.712 | .139 | 26.409 | 1 | .000 | .490 |

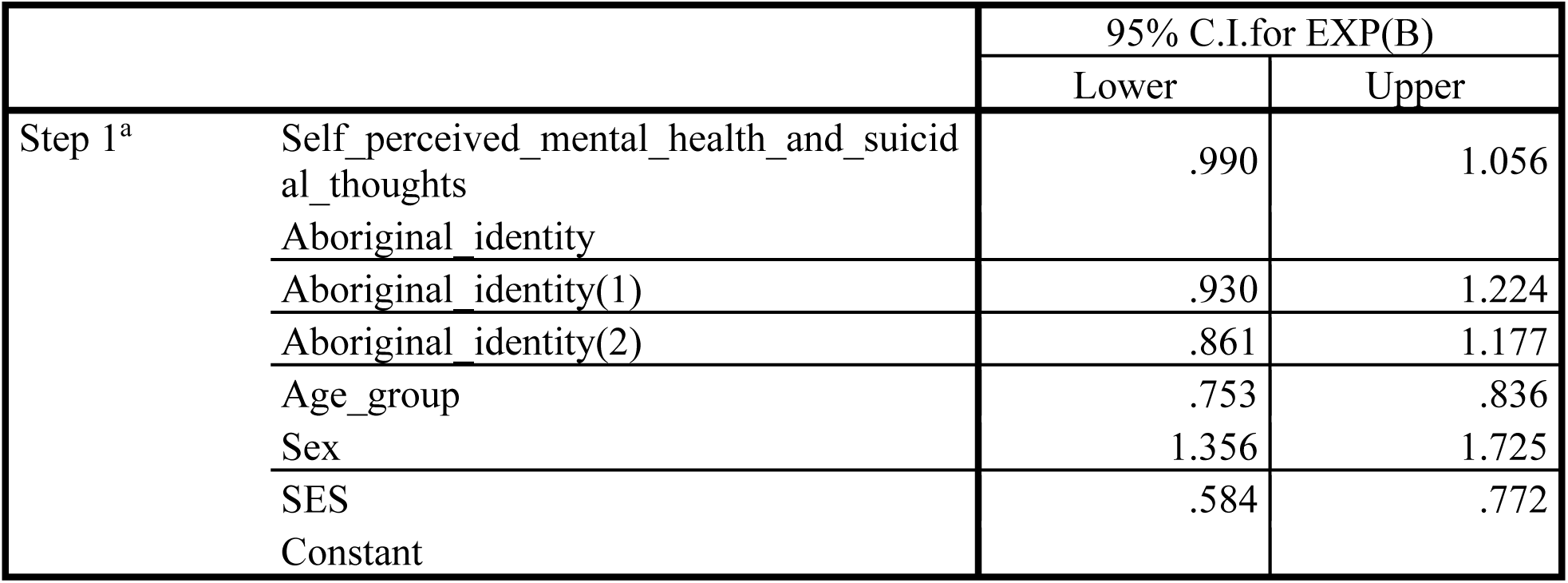
Variables in the Equation.

Variables in the Equation
|  |  | 95% C.I. for EXP(B) |  |
| --- | --- | --- | --- |
|  |  | Lower | Upper |
| Step 1<sup>a</sup> Self_perceived_mental_health_and_suicidal_thoughts |  | .990 | 1.056 |
| Aboriginal_identity |  |  |  |
| Aboriginal_identity(1) |  | .930 | 1.224 |
| Aboriginal_identity(2) |  | .861 | 1.177 |
| Age_group |  | .753 | .836 |
| Sex |  | 1.356 | 1.725 |
| SES |  | .584 | .772 |
| Constant |  |  |  |

a. Variable(s) entered on step 1: Self_perceived_mental_health_and_suicidal_thoughts, Aboriginal_identity, Age_group, Sex, SES.

LOGISTIC REGRESSION VARIABLES Suicidal_Thoughts

/METHOD=ENTER Self_perceived_mental_health_and_suicidal_thoughts

Aboriginal_identity Age_group Sex SES

/CONTRAST (Aboriginal_identity)=Indicator

/CONTRAST (Age_group)=Indicator

/CONTRAST (Sex)=Indicator

/PRINT=GOODFIT CI(95)

/CRITERIA=PIN(0.05) POUT(0.10) ITERATE(20) CUT(0.5).

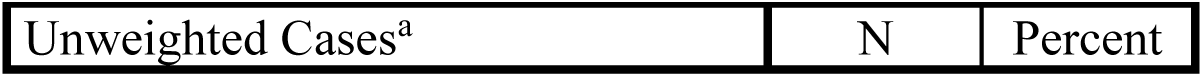
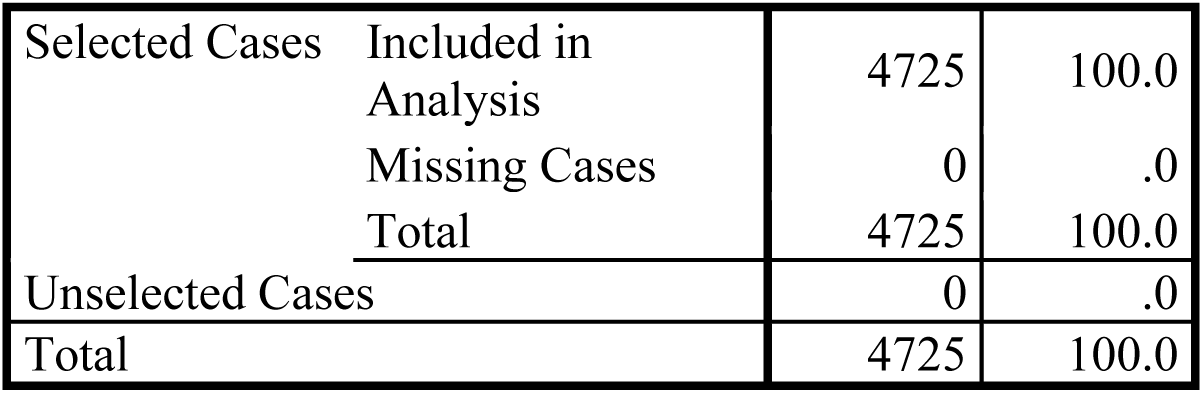
Case Processing Summary.

Case Processing Summary
| Unweighted Cases<sup>a</sup> | N | Percent |
| --- | --- | --- |

|  |  |  |  |
| --- | --- | --- | --- |
| Selected Cases | Included in Analysis | 4725 | 100.0 |
|  | Missing Cases | 0 | .0 |
|  | Total | 4725 | 100.0 |
| Unselected Cases |  | 0 | .0 |
| Total |  | 4725 | 100.0 |

a. If weight is in effect, see classification table for the total number of cases.

Dependent Variable Encoding.

Dependent Variable Encoding
| Original Value | Internal Value |
| --- | --- |
| No | 0 |
| Yes | 1 |

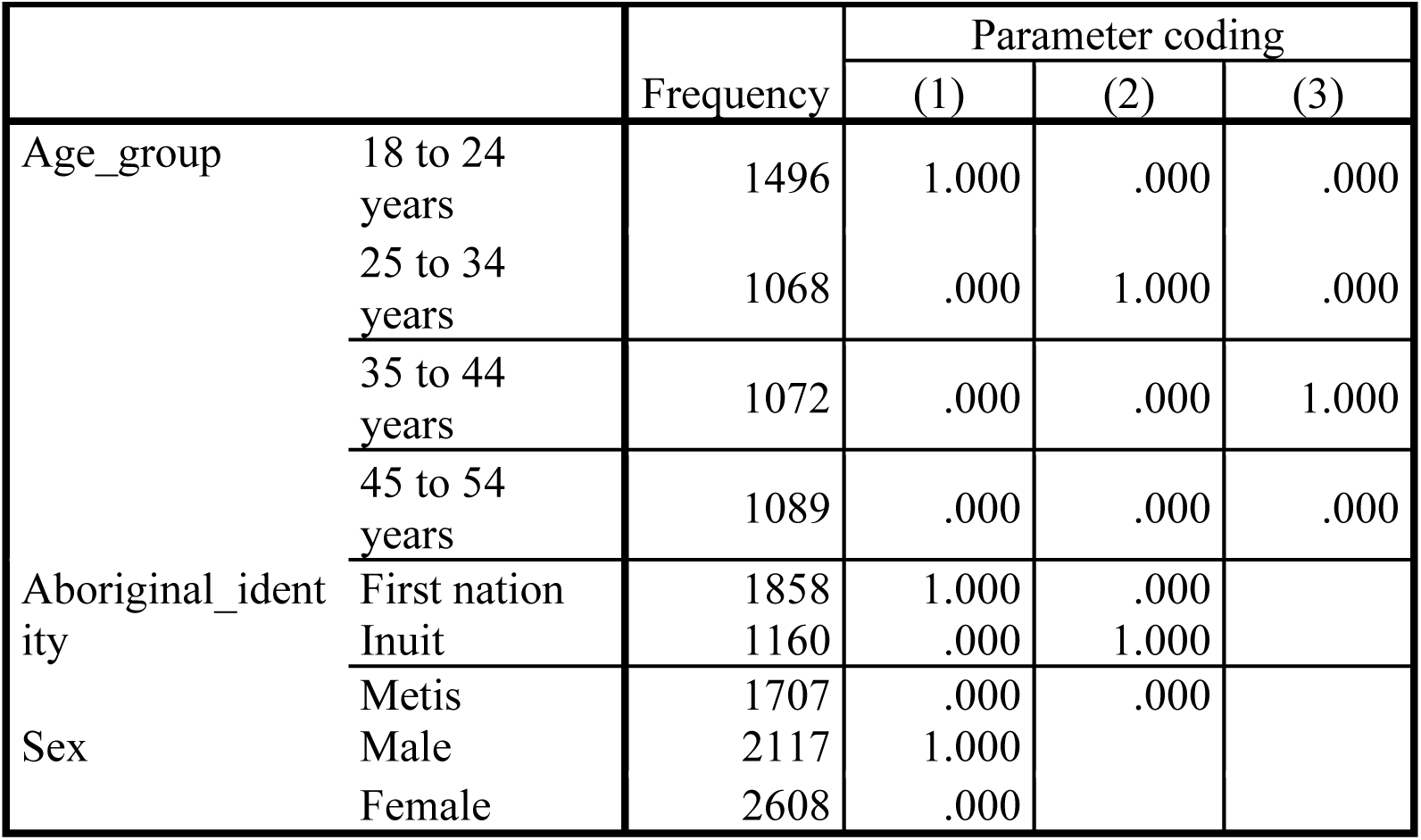
Categorical Variables Codings.

Categorical Variables Codings
|  |  | Frequency | Parameter coding |  |  |
| --- | --- | --- | --- | --- | --- |
|  |  |  | (1) | (2) | (3) |
| Age_group | 18 to 24 years | 1496 | 1.000 | .000 | .000 |
|  | 25 to 34 years | 1068 | .000 | 1.000 | .000 |
|  | 35 to 44 years | 1072 | .000 | .000 | 1.000 |
|  | 45 to 54 years | 1089 | .000 | .000 | .000 |
| Aboriginal_identity | First nation | 1858 | 1.000 | .000 |  |
|  | Inuit | 1160 | .000 | 1.000 |  |
|  | Metis | 1707 | .000 | .000 |  |
| Sex | Male | 2117 | 1.000 |  |  |
|  | Female | 2608 | .000 |  |  |

**Block 0: Beginning Block**

Classification Table^a,b^.

Block 0: Beginning Block Classification Table<sup>a,b</sup>
| Observed |  |  | Predicted |  |  |
| --- | --- | --- | --- | --- | --- |
|  |  |  | Suicidal_Thoughts |  | Percentage Correct |
|  |  |  | No | Yes |  |
| Step 0 | Suicidal_Thoughts | No | 2872 | 0 | 100.0 |
|  |  | Yes | 1853 | 0 | .0 |
| Overall Percentage |  |  |  |  | 60.8 |

a. Constant is included in the model.
b. The cut value is .500

Variables in the Equation.

Variables in the Equation
|  | B | S.E. | Wald | df | Sig. | Exp(B) |
| --- | --- | --- | --- | --- | --- | --- |
| Step 0 Constant | -.438 | .030 | 216.276 | 1 | .000 | .645 |

Variables not in the Equation.

Variables not in the Equation
|  | Score | df | Sig. |
| --- | --- | --- | --- |
| Step 0 Variables Self_perceived_mental_health_and_suicidal_thoughts | 2.822 | 1 | .093 |
| Aboriginal_identity | 1.360 | 2 | .507 |
| Aboriginal_identity(1) | .259 | 1 | .611 |
| Aboriginal_identity(2) | .487 | 1 | .485 |
| Age_group | 113.907 | 3 | .000 |
| Age_group(1) | 113.503 | 1 | .000 |
| Age_group(2) | 11.614 | 1 | .001 |
| Age_group(3) | 19.713 | 1 | .000 |
| Sex(1) | 44.412 | 1 | .000 |
| SES | 38.549 | 1 | .000 |
| Overall Statistics | 196.893 | 8 | .000 |

**Block 1: Method = Enter**

Omnibus Tests of Model Coefficients.

Block 1: Method = Enter Omnibus Tests of Model Coefficients
|  | Chi-square | df | Sig. |
| --- | --- | --- | --- |
| Step 1 Step | 208.362 | 8 | .000 |
| Block | 208.362 | 8 | .000 |
| Model | 208.362 | 8 | .000 |

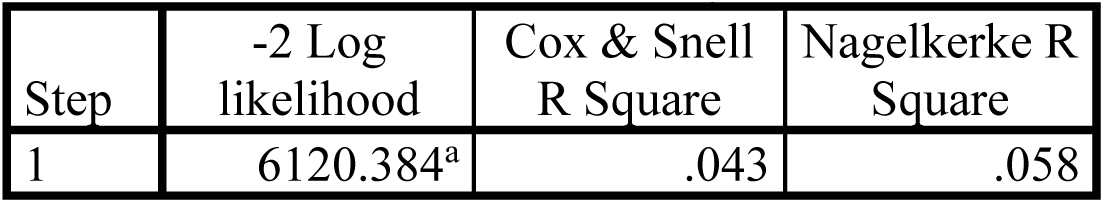
Model Summary.

Model Summary
| Step | -2 Log likelihood | Cox & Snell R Square | Nagelkerke R Square |
| --- | --- | --- | --- |
| 1 | 6120.384<sup>a</sup> | .043 | .058 |

a. Estimation terminated at iteration number 5 because parameter estimates changed by less than .001.

Hosmer and Lemeshow Test.

Hosmer and Lemeshow Test
| Step | Chi-square | df | Sig. |
| --- | --- | --- | --- |
| 1 | 65.353 | 8 | .000 |

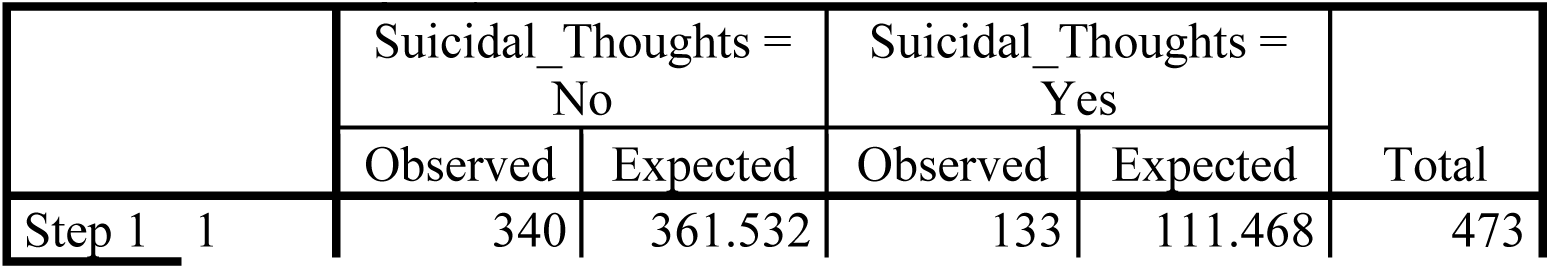
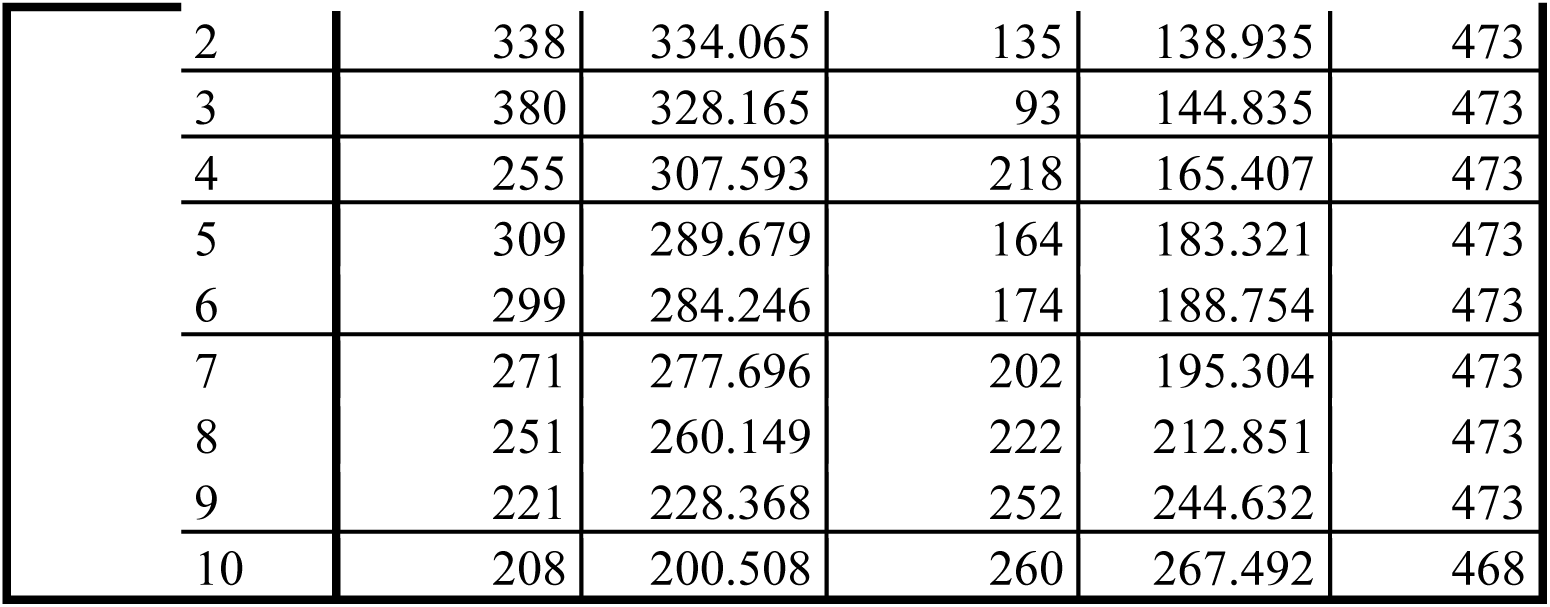
Contingency Table for Hosmer and Lemeshow Test.

Contingency Table for Hosmer and Lemeshow Test
|  |  | Suicidal_Thoughts = No |  | Suicidal_Thoughts = Yes |  | Total |
| --- | --- | --- | --- | --- | --- | --- |
|  |  | Observed | Expected | Observed | Expected |  |
| Step 1 | 1 | 340 | 361.532 | 133 | 111.468 | 473 |

|  |  |  |  |  |  |
| --- | --- | --- | --- | --- | --- |
| 2 | 338 | 334.065 | 135 | 138.935 | 473 |
| 3 | 380 | 328.165 | 93 | 144.835 | 473 |
| 4 | 255 | 307.593 | 218 | 165.407 | 473 |
| 5 | 309 | 289.679 | 164 | 183.321 | 473 |
| 6 | 299 | 284.246 | 174 | 188.754 | 473 |
| 7 | 271 | 277.696 | 202 | 195.304 | 473 |
| 8 | 251 | 260.149 | 222 | 212.851 | 473 |
| 9 | 221 | 228.368 | 252 | 244.632 | 473 |
| 10 | 208 | 200.508 | 260 | 267.492 | 468 |

Classification Table^a^.

**Classification Table<sup>a</sup>**
| Observed |  |  | Predicted |  |  |
| --- | --- | --- | --- | --- | --- |
|  |  |  | Suicidal_Thoughts |  | Percentage Correct |
|  |  |  | No | Yes |  |
| Step 1 | Suicidal_Thought | No | 2521 | 351 | 87.8 |
|  | s | Yes | 1446 | 407 | 22.0 |
|  | Overall Percentage |  |  |  | 62.0 |

a. The cut value is .500

Variables in the Equation.

**Variables in the Equation**
|  | B | S.E. | Wald | df | Sig. | Exp(B) |
| --- | --- | --- | --- | --- | --- | --- |
| Step 1<sup>a</sup> Self_perceived_mental_health_and_suicidal_thoughts | .019 | .016 | 1.337 | 1 | .248 | 1.019 |
| Aboriginal_identity |  |  | .938 | 2 | .626 |  |
| Aboriginal_identity(1) | .067 | .070 | .895 | 1 | .344 | 1.069 |
| Aboriginal_identity(2) | .020 | .080 | .064 | 1 | .800 | 1.020 |
| Age_group |  |  | 109.713 | 3 | .000 |  |
| Age_group(1) | .670 | .083 | 64.896 | 1 | .000 | 1.954 |
| Age_group(2) | .032 | .092 | .121 | 1 | .728 | 1.032 |
| Age_group(3) | -.043 | .092 | .215 | 1 | .643 | .958 |
| Sex(1) | -.434 | .062 | 49.454 | 1 | .000 | .648 |
| SES | -.389 | .071 | 29.894 | 1 | .000 | .678 |
| Constant | -.609 | .101 | 36.629 | 1 | .000 | .544 |

Variables in the Equation.

**Variables in the Equation**
|  |  | 95% C.I.for EXP(B) |  |
| --- | --- | --- | --- |
|  |  | Lower | Upper |
| Step 1<sup>a</sup> | Self_perceived_mental_health_and_suicidal_thoughts | .987 | 1.053 |
|  | Aboriginal_identity |  |  |
|  | Aboriginal_identity(1) | .931 | 1.227 |
|  | Aboriginal_identity(2) | .872 | 1.194 |
|  | Age_group |  |  |
|  | Age_group(1) | 1.660 | 2.300 |
|  | Age_group(2) | .863 | 1.235 |
|  | Age_group(3) | .800 | 1.147 |
|  | Sex(1) | .574 | .731 |
|  | SES | .589 | .779 |
|  | Constant |  |  |

a. Variable(s) entered on step 1: Self_perceived_mental_health_and_suicidal_thoughts, Aboriginal_identity, Age_group, Sex, SES.

FREQUENCIES VARIABLES=Aboriginal_identity Age_group Sex Self_perceived_mental_health_and_suicidal_thoughts Suicidal_Thoughts

/ORDER=ANALYSIS.

**Frequencies**

Statistics.

Statistics
|  |  | Aboriginal_i<br>dentity | Age_group | Sex | Self_perceive<br>d_mental_he<br>alth_and_suic<br>idal_thoughts | Suicidal_Tho<br>ughts |
| --- | --- | --- | --- | --- | --- | --- |
| N | Valid | 4725 | 4725 | 4725 | 4725 | 4725 |
|  | Missing | 0 | 0 | 0 | 0 | 0 |

**Frequency Table**

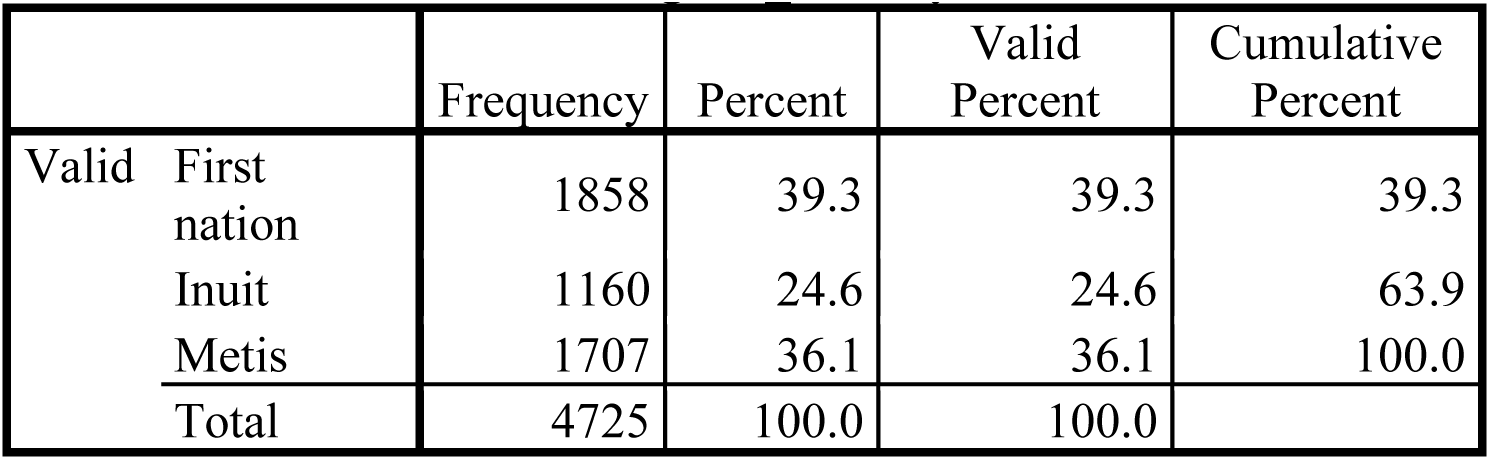
Aboriginal_identity.

Age_group.

Age\_group
|  |  | Frequency | Percent | Valid<br>Percent | Cumulative<br>Percent |
| --- | --- | --- | --- | --- | --- |
| Valid | 18 to 24 years | 1496 | 31.7 | 31.7 | 31.7 |
|  | 25 to 34 years | 1068 | 22.6 | 22.6 | 54.3 |
|  | 35 to 44 years | 1072 | 22.7 | 22.7 | 77.0 |
|  | 45 to 54 years | 1089 | 23.0 | 23.0 | 100.0 |
|  | Total | 4725 | 100.0 | 100.0 |  |

Sex.

Sex
|  | Frequency | Percent | Valid Percent | Cumulative Percent |
| --- | --- | --- | --- | --- |
| Valid Male | 2117 | 44.8 | 44.8 | 44.8 |
| Female | 2608 | 55.2 | 55.2 | 100.0 |
| Total | 4725 | 100.0 | 100.0 |  |

Self_perceived_mental_health_and_suicidal_thoughts.

Self\_perceived\_mental\_health\_and\_suicidal\_thoughts
|  | Frequency | Percent | Valid Percent | Cumulative Percent |
| --- | --- | --- | --- | --- |
| Valid Ever seriously contemplated suicide | 632 | 13.4 | 13.4 | 13.4 |
| Excellent or very good self-perceived mental health | 968 | 20.5 | 20.5 | 33.9 |
| Fair or poor self-perceived mental health | 489 | 10.3 | 10.3 | 44.2 |
| Good self-perceived mental health | 886 | 18.8 | 18.8 | 63.0 |
| Never seriously contemplated suicide | 1018 | 21.5 | 21.5 | 84.5 |
| Seriously contemplated suicide in the past 12 months | 232 | 4.9 | 4.9 | 89.4 |
| Seriously contemplated suicide but not in the past 12 months | 500 | 10.6 | 10.6 | 100.0 |
| Total | 4725 | 100.0 | 100.0 |  |

Suicidal_Thoughts.

Suicidal\_Thoughts
|  | Frequency | Percent | Valid Percent | Cumulative Percent |
| --- | --- | --- | --- | --- |
| Valid No | 2872 | 60.8 | 60.8 | 60.8 |
| Yes | 1853 | 39.2 | 39.2 | 100.0 |
| Total | 4725 | 100.0 | 100.0 |  |

